# Sensitivity, throughput, and cost analysis of concentration methods for multi-target pathogen wastewater monitoring

**DOI:** 10.1101/2025.05.12.25327458

**Authors:** Jingjing Wu, Michael X. Wang, Todd J. Treangen, Katherine Ensor, Loren Hopkins, Lauren B. Stadler

## Abstract

Wastewater-based epidemiology is an efficient method for monitoring the transmission of diverse pathogens in communities. Standard wastewater surveillance workflows typically involve wastewater concentration, nucleic acid extraction, and pathogen quantification. While various concentration methods are used, most comparisons of concentration methods have focused primarily on SARS-CoV-2, highlighting the need for further research to guide method selection for monitoring a suite of diverse pathogens. In this study, a head-to-head comparison of six different concentration methods was performed, including direct extraction (with and without bead beating), electronegative (HA) filtration, solids concentration, and magnetic bead-based concentration (using Nanotrap® particles; with and without bead beating). Methods were assessed for sensitivity, inhibitor removal, and recovery rates of fourteen microorganisms, including viruses, bacteria, and fungal pathogens. The cost of each method was also estimated. Results showed that the concentration method selection significantly impacts the sensitivity and economic costs of the wastewater monitoring workflow. Based on the results, a concentration approach that combines HA filtration and solids concentration is recommended to optimize detection across various pathogens. This study provides data-driven insights to enhance the reliability and cost-effectiveness of wastewater surveillance systems that can support public health responses for a broad range of diseases.

**Synopsis:** Six concentration methods were compared in terms of sensitivity and cost for the detection of 14 diverse pathogens in wastewater.

## Introduction

Wastewater-based epidemiology has been widely used as a resource-efficient and cost-effective method to monitor infectious diseases. Wastewater is a rich source of information on community health and contains many human pathogens that can be quantified to reveal their presence and epidemiology in communities. Wastewater monitoring programs greatly expanded in response to the SARS-CoV-2 pandemic and have since grown to include many other pathogen targets. Wastewater surveillance has been applied for many respiratory viruses, including SARS-CoV-2, influenza A, influenza B, and respiratory syncytial virus (RSV), as well as a broader range of microorganisms, such as enteric viruses, fungal and bacterial pathogens such as *Candida auris* and *Salmonella* ^1–16^. Sustaining a global wastewater monitoring system will require demonstrating the benefits of the surveillance system, including understanding which targets to prioritize and tradeoffs associated with expanding the number and types of targets as part of a routine monitoring program.

Methods for detecting and quantifying pathogen targets in wastewater typically include three steps: (1) a concentration step as pathogen targets are present at very low levels in wastewater; (2) nucleic acid extraction and purification; and (3) quantification that typically involves PCR amplification of target nucleic acids. A variety of different wastewater concentration methods are in use, within them, centrifugation- and filtration-based concentration methods were among the most commonly used methods for wastewater monitoring ^17–21^. Additionally, many labs are adopting more automated wastewater concentration methods, such as those that use the magnetic bead capture method (e.g., using Nanotrap® particles, referred to as Nanotrap method) ^20,22,23^.

Multiple studies have been conducted to compare the impacts of wastewater concentration methods on wastewater monitoring results, including centrifugation, filtration, and Nanotrap methods. However, these studies have largely focused on SARS-CoV-2. For example, Laturner et al. (2021) evaluated direct extraction, electronegative (HA) filtration, polyethylene glycol (PEG) precipitation, and ultrafiltration for detecting SARS-CoV-2 in wastewater, and showed that HA filtration with bead beating was most sensitive and efficient ^21^. Similarly, Ahmed et al. (2020) reported HA filtration as the most efficient method for SARS-CoV-2 monitoring in wastewater compared to centrifugal filtration, PGE precipitation, and ultrafiltration^18^. Several other methods comparison studies have assessed the Nanotrap method for SARS-CoV-2 detection in wastewater; however, these studies reported inconsistent results. For example, a study by Liu et al. (2023) showed that using Nanotrap method yielded higher concentrations of SARS-CoV-2 RNA in wastewater than the HA filtration method ^23^. In contrast, Ahmed et al. (2023) observed higher concentration of SARS-CoV-2 RNA in wastewater samples using HA filtration than the Nanotrap method ^22^.

Few studies have compared wastewater concentration methods for detecting targets other than SARS-CoV-2, or addressed the tradeoffs in sensitivity for different types of targets (e.g., viruses, bacteria, protozoa) using a single method for concentration as part of a routine wastewater monitoring program. Zheng et al. (2023) reported higher concentrations of influenza A and B when assaying the solids fraction of influent wastewater after low-speed centrifugation as compared to the liquid fraction using PEG precipitation ^24^. Ahmed et al. (2021) observed higher concentrations of adenovirus in wastewater using HA filtration of the influent as compared to concentrations measured using the wastewater solids ^25^. Hachimi et al. (2024) evaluated multiple wastewater concentration methods for detecting *Cryptosporidium* in wastewater and observed the highest recovery when assaying the solid fraction of influent after centrifugation compared to other methods, including HA filtration and the Nanotrap method ^20^. Importantly, the targets varied across each of these prior studies comparing different concentration methods, which limits the comparability of the results for selecting a concentration method for targeting a suite of diverse pathogens.

In this study, we performed a head-to-head comparison of six different concentration methods in terms of sensitivity, inhibitor removal, and recovery rates of fourteen different microorganisms in wastewater, including viruses, bacteria, and fungal pathogens. The concentration methods included in the comparison were: 1) the direct extraction of nucleic acids from wastewater; 2) the direct extraction of nucleic acids from wastewater with a bead-beating step; 3) HA filtration of the liquid fraction of wastewater with a bead-beating step (HA filtration method); 4) concentration of the solids fraction of wastewater via centrifugation with a bead-beating step (solids method); 5) magnetic bead capture with Nanotrap® particles (Nanotrap method); and 6) the Nanotrap method with a bead-beating step. In addition to comparing the performance of these methods for a diverse suite of targets, we estimated and compared the economic costs of these concentration methods. Based on these results, we provide data-driven guidance on the selection of concentration methods for wastewater monitoring of diverse microorganisms beyond SARS-CoV-2 and respiratory viruses.

## Methods

### Wastewater sample collection and preparation

Composite influent wastewater samples were collected from wastewater treatment plants (WWTPs) in Houston on December 4, 2023, using a refrigerated time-weighted autosampler as described previously ^26^. Wastewater samples were transported to Rice University on ice and stored at 4°C for less than 48 hours before processing. Fifty mL of influent from 10 different Houston WWTPs were combined in a 500 mL bottle and inverted multiple times to mix before being aliquoted into two 250 mL bottles (bottle A and bottle B).

Pathogen standards (Table 1) were diluted, mixed, and spiked into one of the 250 mL bottles of wastewater (bottle A) prior to concentration (details described in SI 1.2). The selection of pathogens to spike was based on baseline concentrations of each pathogen in Houston wastewater from our routine monitoring system (data not shown). For pathogens that were spiked into the wastewater sample, the target final concentration for each was between 10^4.72^ and 10^7.25^ copies/L. The remaining 250 mL of the mixed wastewater (bottle B) was processed without any spike-ins, undergoing wastewater concentration and nucleic acid extraction.

**Table 1.** Pathogen targets, multiplexing information, and details of pathogen standards spiked into the wastewater sample. For spiked pathogens, the target microbial concentrations were measured in both raw and spiked wastewater samples. For the unspiked pathogens, target microbial concentrations were measured only in raw influent wastewater samples.

| Plate code | Virus/Target | Fluorophore | Spike-in standard |
| --- | --- | --- | --- |
| Respiratory viruses | Influenza A | FAM | ATCC, VR-1894 |
|  | Human respiratory syncytial virus (RSV) | SUN | ATCC, VR-1803 |
|  | SARS-CoV-2 | Cy5.5 | VR-3347HK |
|  | Influenza B | ROX | ATCC, VR-1804 |
| Pathogenic microorganisms | *Cryptosporidium* | FAM | University of Arizona, Lot #230719 |
|  | *Candida auris* | SUN | ATCC, MYA-5001 |
|  | *Salmonella enterica* | Cy5 | ATCC, 14028 |
|  | *Campylobacter jejuni* | Cy5.5 | ATCC, 33560 |
| Enteric viruses, group 1 | Sapovirus | FAM | Not spiked |
|  | Astrovirus | SUN | Not spiked |
|  | Rotavirus A | Cy5 | Not spiked |
|  | Adenovirus 41 | Cy5.5 | Not spiked |
| Enteric viruses, group 2 | Norovirus GI | FAM | Not spiked |
|  | Norovirus GII | SUN | Not spiked |

### Concentration of pathogen targets in wastewater

We compared the concentrations of a suite of microbial targets (Table 1) in wastewater samples using six concentration methods (Figure 1). Each concentration method was performed in triplicate using wastewater spiked with pathogen standards (bottle A) and in triplicate using raw influent wastewater (bottle B). The concentrates were stored at 4°C for less than 4 hours before nucleic acid extraction.

**Figure 1.**
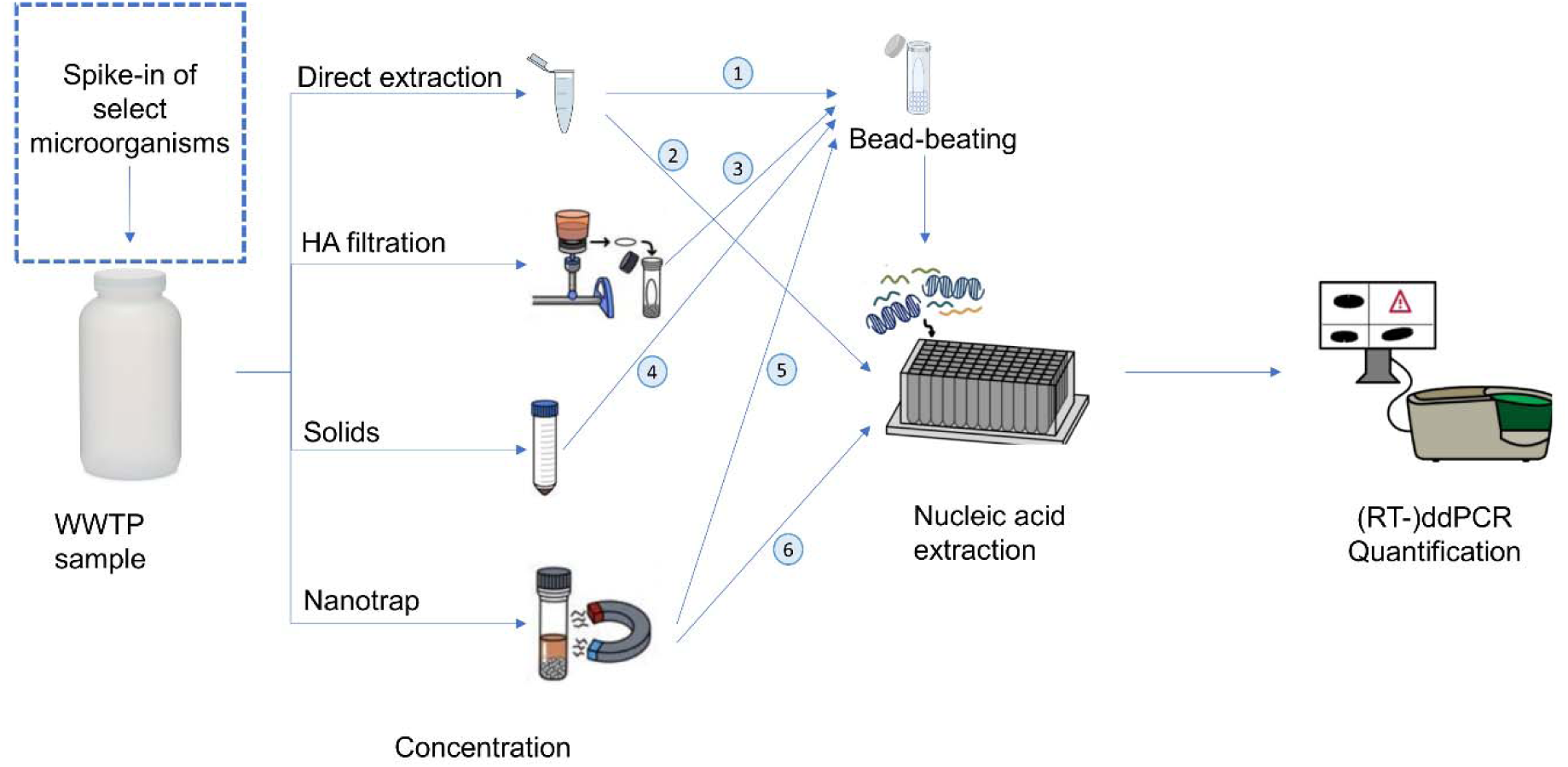
Overview of the concentration methods and quantification workflow. The wastewater samples were processed using six different concentration methods: 1) direct extraction with bead beating, 2) direct extraction, 3) solids fraction with bead beating, 4) HA filtration of supernatant with bead beating, 5) Nanotrap beads with bead beating, and 6) Nanotrap beads without bead beating.

**Figure 2.**
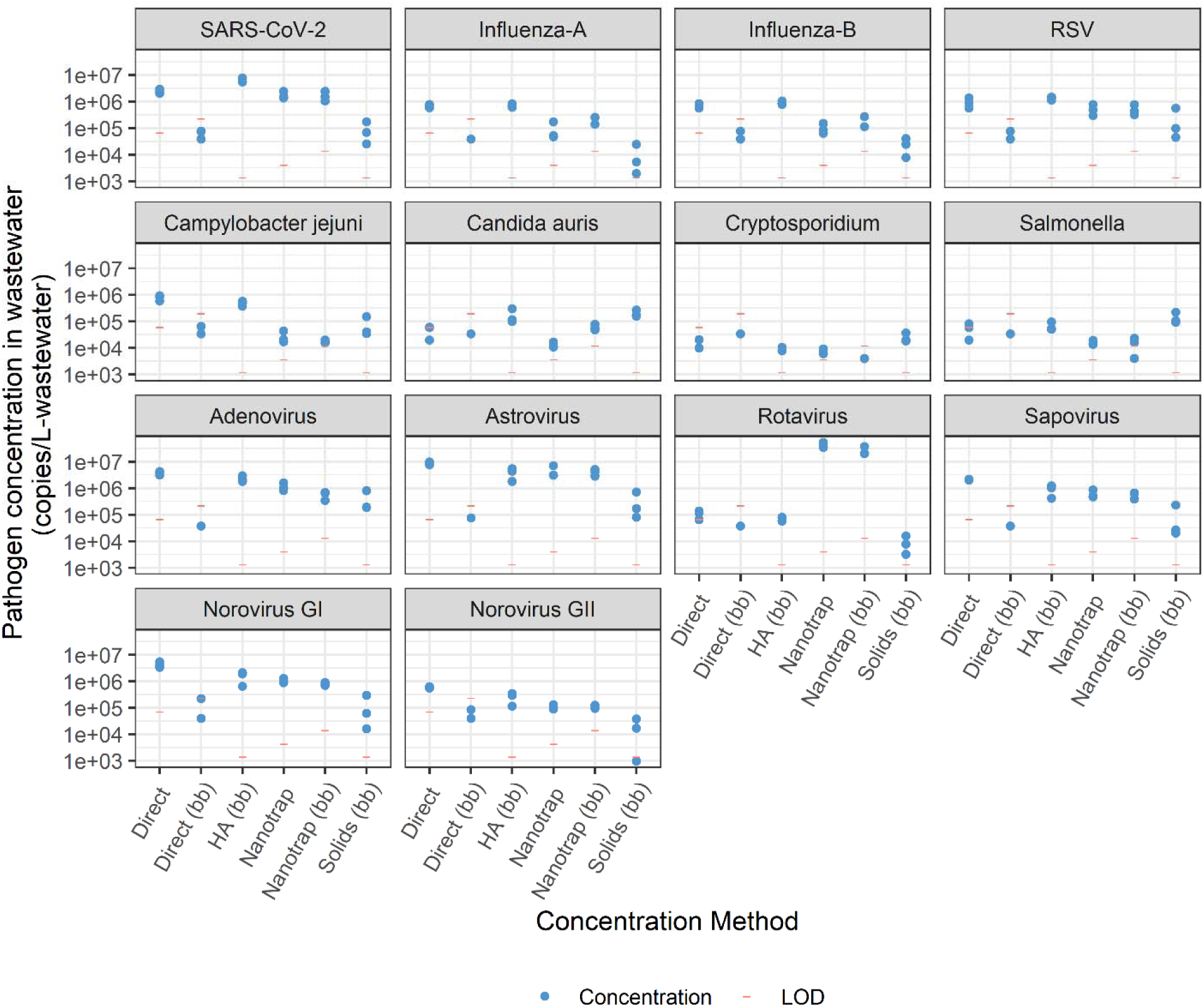
Impact of wastewater concentration method on pathogen nucleic acid concentrations in wastewater samples. Concentrations of SARS-CoV-2, influenza A, influenza B, RSV, *Campylobacter jejuni*, *Candida auris*, *Cryptosporidium*, and *Salmonella* measured in wastewater spiked with a mixture of pathogens (Table 1), and concentrations of adenovirus, astrovirus, rotavirus, sapovirus, norovirus GI, and norovirus GII measured in raw wastewater samples.

#### Direct extraction

One mL of wastewater was aliquoted into 1.5 mL tubes (022431021, Eppendorf) and centrifuged for 5 minutes at 17,000 g and 4°C. 300 µL of the supernatant was moved to a filled bead-beating tube containing 0.1 mm diameter glass beads and 700 µL of lysis buffer from the Chemagic^TM^ Prime Viral DNA/RNA 300 Kit H96 (CMG-1433, PerkinElmer) for bead beating (detailed in SI 1.2) and nucleic acid extraction. Another 300 µL of the supernatant from the 1.5 mL tube was aspirated without disturbing the pellet and used directly for nucleic acid extraction.

#### HA filtration and solids method

Fifty mL of wastewater sample was aliquoted into 50 mL tubes (21008-242, VWR). The wastewater liquid and solid fractions were separated by centrifuging the tube for 20 minutes at 4,100 g and 4°C. After centrifugation, the supernatant was concentrated by passing through an Electronegative Microbiological Analysis Membrane HA Filter (HAWG047S6, Millipore Sigma) as described previously ^21,26^. For the solids fraction, the pellet that remained in the tube was resuspended with 1 mL of lysis buffer from the Chemagic^TM^ Prime Viral DNA/RNA 300 Kit H96 (CMG-1433, PerkinElmer) and moved to a filled bead-beating tube containing 0.1 mm diameter glass beads before bead beating and nucleic acid extraction.

#### Nanotrap method

Ten mL of wastewater sample was aliquoted into a 15 mL Falcon tube (352096, Fisher Scientific) and concentrated into 600 µL of lysis buffer from the ChemagicTM Prime Viral DNA/RNA 300 Kit H96 (CMG-1433, PerkinElmer) using the Nanotrap^®^ Microbiome A and B particles (44202 and 65202, Ceres Nanosciences), following the protocol from Ceres Nanosciences, Inc (Manassas, VA; details described in SI 1.2). 300 µL of concentrate was transferred to a filled bead-beating tube (0.1 mm diameter glass beads and 700 µL of lysis buffer from the ChemagicTM Prime Viral DNA/RNA 300 Kit H96 (CMG-1433, PerkinElmer)) for bead beating (detailed in SI 1.2) and nucleic acid extraction. The remaining 300 µL of concentrate was used directly for nucleic acid extraction.

### Nucleic acid extraction and target nucleic acid quantification

Nucleic acid extraction was performed using ChemagicTM Prime Viral DNA/RNA 300 Kit H96 (Chemagic, CMG-1433, PerkinElmer). The concentrations of target viral RNA were quantified using the one-step RT-ddPCR Advanced Kit for Probes (1864021, Bio-Rad), and the concentrations of target microbial DNA were quantified using the ddPCR^TM^ Supermix for Probes (1863024, Bio-Rad) (details described in SI 1.2). Primer and probe sequences for each assay used in this study, (RT-)ddPCR reaction assay composition, and thermal cycler conditions are provided in SI 1.4. We multiplexed the ddPCR assays for resource efficiency and to reduce the overall cost of quantification, and the assays were multiplexed based on the requirements of the ddPCR kits and the thermal cycler conditions. We followed the EMMI Guidelines ^27^ for quality control; additional information is provided in Table SI.14.

### Comparison of inhibition and recovery rates

We characterized inhibition associated with each concentration method and each target. We evaluated the presence of inhibitors by diluting the nucleic acid extracts 10-fold using nuclease-free water (AM9930, ThermoFisher). We calculated an inhibition factor as the concentration based on 10x diluted extracts divided by the concentration from the undiluted extracts (SI. Eq 2). The data were excluded from analysis when the measured concentrations based on 10x diluted extracts were less than 1.5 times the limit of detection (LOD).

For HA filtration, solids, and Nanotrap (with and without bead beating) methods, we calculated recovery rates (SI. Eq 1) to evaluate the performance of the concentration, extraction, and quantification workflow for pathogens spiked into wastewater samples. The quantity of each pathogen spiked into the wastewater sample was determined based on the concentration of target in the spike-in solution mixture and the volume of solution spiked into the sample. To measure the microbial concentrations in the spike-in solution, we utilized the ChemagicTM Prime Viral DNA/RNA 300 Kit H96 (Chemagic, CMG-1433, PerkinElmer) and heat lysis (at 95°C for 10 minutes), followed by quantification using (RT-)ddPCR. For the nucleic acid extraction pretreatment method, we processed the spike-in solution with or without a bead-beating step (detailed in SI 1.2). The highest of the three measured concentrations was used to calculate recovery rates (SI 2.2).

### Cost comparison of different wastewater concentration methods

We performed a cost comparison to guide decision-making related to choosing a wastewater concentration method given a set of wastewater surveillance targets. This cost analysis included both capital and operational costs. The capital costs included instrument and equipment costs. The operational costs included the costs of consumables and labor. We assumed duplicate wastewater concentration for each sample. For the Nanotrap method, we assumed a KingFisher Apex Purification System (5400930, Thermo Scientific) would be used for wastewater sample processing. Manual wastewater processing was assumed for all other concentration methods. This analysis excluded the equipment maintenance costs, as they are typically proportional to and thus reflected in the capital costs. All the information for cost analysis was from purchase history, list prices, or lab records. The costs are presented in 2024 U.S. dollars.

### Statistical analysis

Statistical analysis was conducted using RStudio (2024.09.0, R version 4.4.1). We compared the log-transformed microbial concentrations in wastewater (as copies/L-wastewater) measured using different concentration methods. The Shapiro-Wilk Test was used to test the normality of the results. The t-test was applied to assess statistical significance when the data were normally distributed, and the Wilcoxon signed-rank test was used for the significance test for non-normally distributed results.

## Results & Discussion

### Impact of the concentration method on measured concentrations and limits of detection

We first compared the quantitative results of six concentration methods for measuring the concentrations of pathogen targets (listed in Table 1) in wastewater samples (Figure 1). Positive detections were observed for all targets when the wastewater samples were concentrated using the HA filtration, solids, or Nanotrap methods. For respiratory viruses, we observed the highest concentrations of SARS-CoV-2, influenza B, and RSV when the wastewater samples were processed using HA filtration and the second highest measurements with direct extraction. We observed significantly higher SARS-CoV-2 concentrations using HA filtration than direct extraction (t-test, p < 0.01), and no significant difference in concentrations of influenza B and RSV using HA filtration and direct extraction (t-test, p > 0.1). For SARS-CoV-2 detection, our results align with the results from Ahmed et al. (2023, 2020) and Laturner et al. (2021), indicating that HA filtration performed best as compared to other concentration methods, including direct extraction, solids method, and Nanotrap method ^18,21,22^. However, the results were inconsistent with those reported by Liu et al. (2023), which found that the Nanotrap method performed better than HA filtration for SARS-CoV-2 detection in wastewater ^23^. The difference may be due to the composition of the wastewater samples and/or the nucleic acid extraction kit used, which are discussed further below. For influenza A, we observed the highest viral concentrations with direct extract and the second highest concentration with HA filtration, however, the difference was not significant (t-test, p > 0.1). Lower measured concentrations were observed with the Nanotrap method. Using the solids method and direct extraction with bead beating resulted in the lowest observed viral concentrations for all the respiratory viruses in this study.

For the pathogenic microorganism *Campylobacter jejuni*, we observed the highest measured concentrations with direct extraction and HA filtration, and there was no significant difference between the methods (t-test p > 0.05). The measured concentrations were significantly higher than those determined using other concentration methods (t-test p < 0.05). The two highest concentrations observed for the other target pathogenic microorganisms were with HA filtration and the solids method. No significant differences in concentrations were observed for *Candida auris* and *Salmonella enterica* between HA filtration and the solids method (t-test, p > 0.1), while the *Cryptosporidium* concentration was significantly higher using the solids method than HA filtration. Similar results were observed by Hachimi et al. (2024), and their study also indicated that the solids method had better performance in detecting *Cryptosporidium* compared to HA filtration and the Nanotrap method ^20^.

For the enteric viruses adenovirus, sapovirus, norovirus GI, and norovirus GII, the highest concentrations were measured using direct extraction and HA filtration, and no significant difference was observed between the methods (t-test, p > 0.05). Lower levels of target viral concentrations were observed when the wastewater samples were concentrated using other methods. The highest astrovirus concentrations were measured using direct extraction, HA filtration, and the Nanotrap method, and no significant difference was observed between these methods (t-test, p > 0.1). The Nanotrap method yielded the highest rotavirus concentration in wastewater, and all other methods resulted in significantly lower measured concentrations (t-test, p < 0.05).

Each concentration method also has a different theoretical LOD, which impacts its sensitivity for pathogen quantification. The theoretical LOD for each method was determined assuming a constant minimum number of positive droplets in the ddPCR reaction and a method-specific concentration factor (SI 1.6, theoretical LOD for each method presented in Table SI.13). The concentration factor is a key metric for evaluating the performance of concentration methods in terms of sensitivity, as a higher concentration factor leads to higher target DNA or RNA concentrations in the (RT-)ddPCR extract, assuming equal microbial levels in the wastewater samples. In this study, the HA filtration and the solids methods had the highest concentration factor of 300, leading to the lowest theoretical LOD of 1,431 copies/L-wastewater. The concentration for the Nanotrap was 100, and thus the theoretical LOD increased to 4,295 copies/L-wastewater. The direct extraction method had a theoretical LOD of 71,586 copies/L-wastewater and yielded extracts that were 50-fold less concentrated compared to those processed using HA filtration or the solids method. The addition of bead beating to the direct extraction or Nanotrap workflow resulted in a 3.3-fold increase in the theoretical LOD. In addition to the recovery of target pathogens in wastewater, the concentration factor and theoretical LOD are critical to the sensitivity of a concentration method. High theoretical LODs may result in insufficient sensitivity for detecting pathogens present at low levels in wastewater samples.

In sum, our results indicate that the selection of concentration method significantly impacted the sensitivity and concentrations of the suite of pathogen targets in wastewater. HA filtration performed best in terms of measured concentrations while also having a relatively low theoretical LOD for *Campylobacter jejuni*, adenovirus, and all respiratory viruses included in this study (SARS-CoV-2, influenza A, influenza B, and RSV). Using the solids method provided the most sensitive detection for *Candida auris*, *Cryptosporidium*, and *Salmonella*. Processing wastewater with either HA filtration or Nanotrap method can reliably detect astrovirus, sapovirus, norovirus GI, and norovirus GII. The Nanotrap method provided the most sensitive detection for rotavirus. In addition, our results suggest that bead-beating does not improve sensitivity using direct extraction or the Nanotrap method, as it did not significantly improve the recovery of pathogen targets in wastewater and it increased the method’s theoretical LOD.

### Impact of concentration method on inhibition and recovery rates of target microorganisms in wastewater

We evaluated the impact of inhibition on pathogen detection and quantification for each concentration method (Figure 3). We used the target concentration measured in the 10-fold diluted nucleic acid extract divided by that in the undiluted extract to calculate an inhibition factor (SI Eq. 2). An inhibition factor threshold of 1.41, as recommended by Cao et al. 2012 and Kang et al. 2019 ^28,29^, was used to indicate the presence of inhibition; inhibition factors above this threshold were considered indicative of inhibition. Using the solids method, substantial inhibition was observed: inhibition factors were above the threshold for SARS-CoV-2 (2.89 ± 1.38), influenza A (2.98 ± 1.00), influenza B (1.79 ± 2.44), astrovirus (2.21 ± 0.96), norovirus GI (2.79 ± 1.05), and norovirus GII (4.16 ± 2.15). For the Nanotrap method, inhibition was observed for the targets SARS-CoV-2 (2.88 ± 1.49), influenza A (4.59 ± 2.21), influenza B (1.55 ± 0.29), RSV (1.46 ± 0.38), astrovirus (2.13 ± 0.27), norovirus GI (2.63 ± 0.59), and norovirus GII (2.70 ± 0.17). Slight inhibition was observed for SARS-CoV-2 with direct extraction (1.50 ± 1.05).

**Figure 3.**
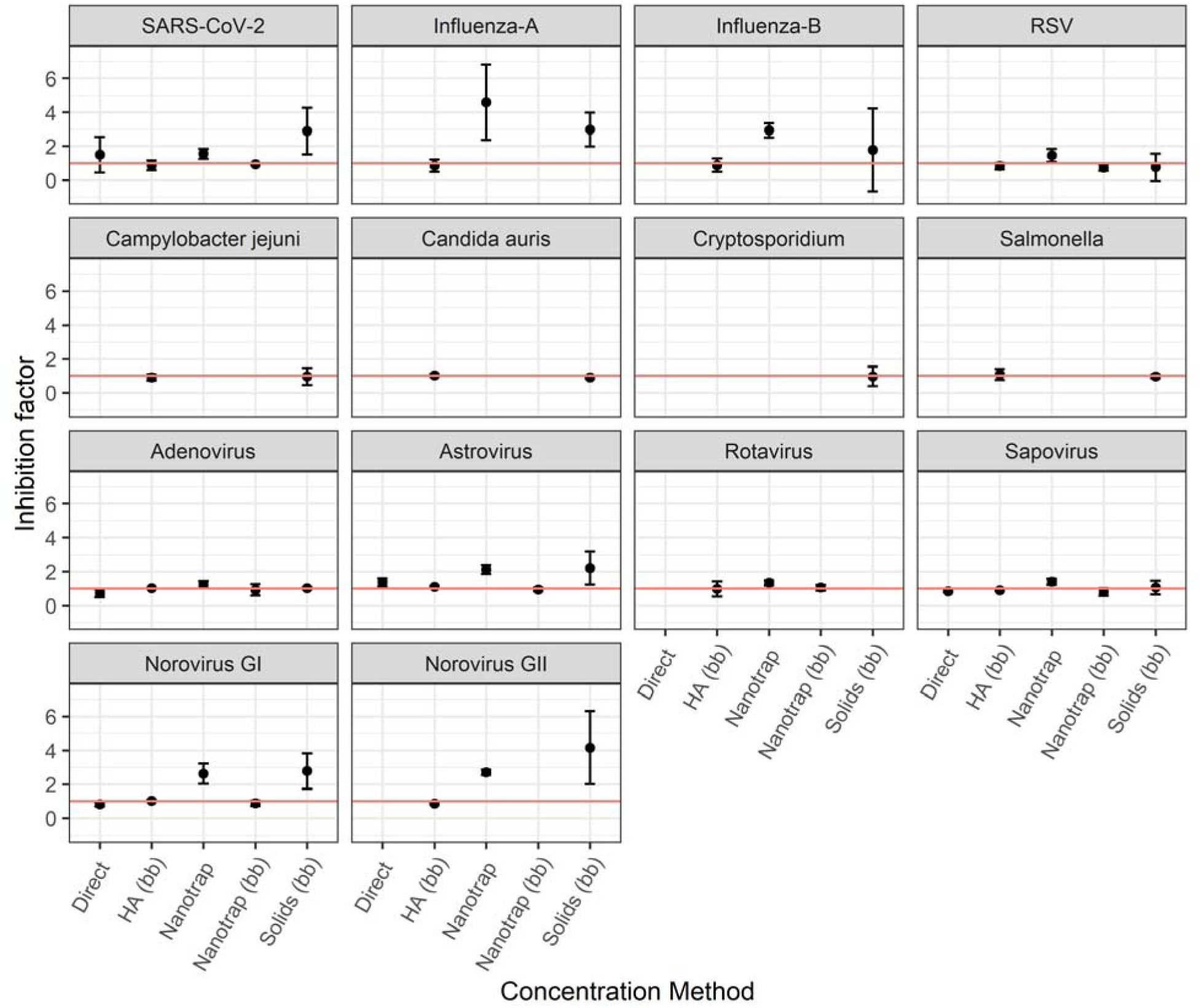
Impact of wastewater concentration method on inhibitors that impact quantification of target pathogens in wastewater. The inhibition factor was calculated using concentrations measured using 10-fold diluted nucleic acid extracts divided by the concentrations measured using undiluted nucleic acid extracts. The red line indicates an inhibition factor of 1. The inhibition factor was reported when the measured concentration of the 10-fold diluted nucleic acid extract was greater than 1.5 times the limit of detection.

In addition to the inhibition, we compared the recovery rates of the spiked pathogen targets when the wastewater samples were concentrated using HA filtration, the Nanotrap method (with and without the bead-beating step), and the solids method (Figure 4). For the respiratory viruses, we observed the highest recovery rates when the wastewater samples were processed using HA filtration. The recovery was above 25% for SARS-CoV-2 and RSV (SARS-CoV-2: 25.8 ± 4.5%; RSV: 26.2 ± 3.4%). Lower recovery was observed for influenza A and B: 12.8 ± 2.2% for influenza A and 16.0 ± 2.5% for influenza B. The recovery was 10.2 ± 4.6% and 10.5 ± 4.9% when the RSV concentrations were quantified with the Nanotrap method with and without the bead-beating step, respectively. For all the other respiratory viruses and concentration methods, recovery rates were below 10%.

**Figure 4.**
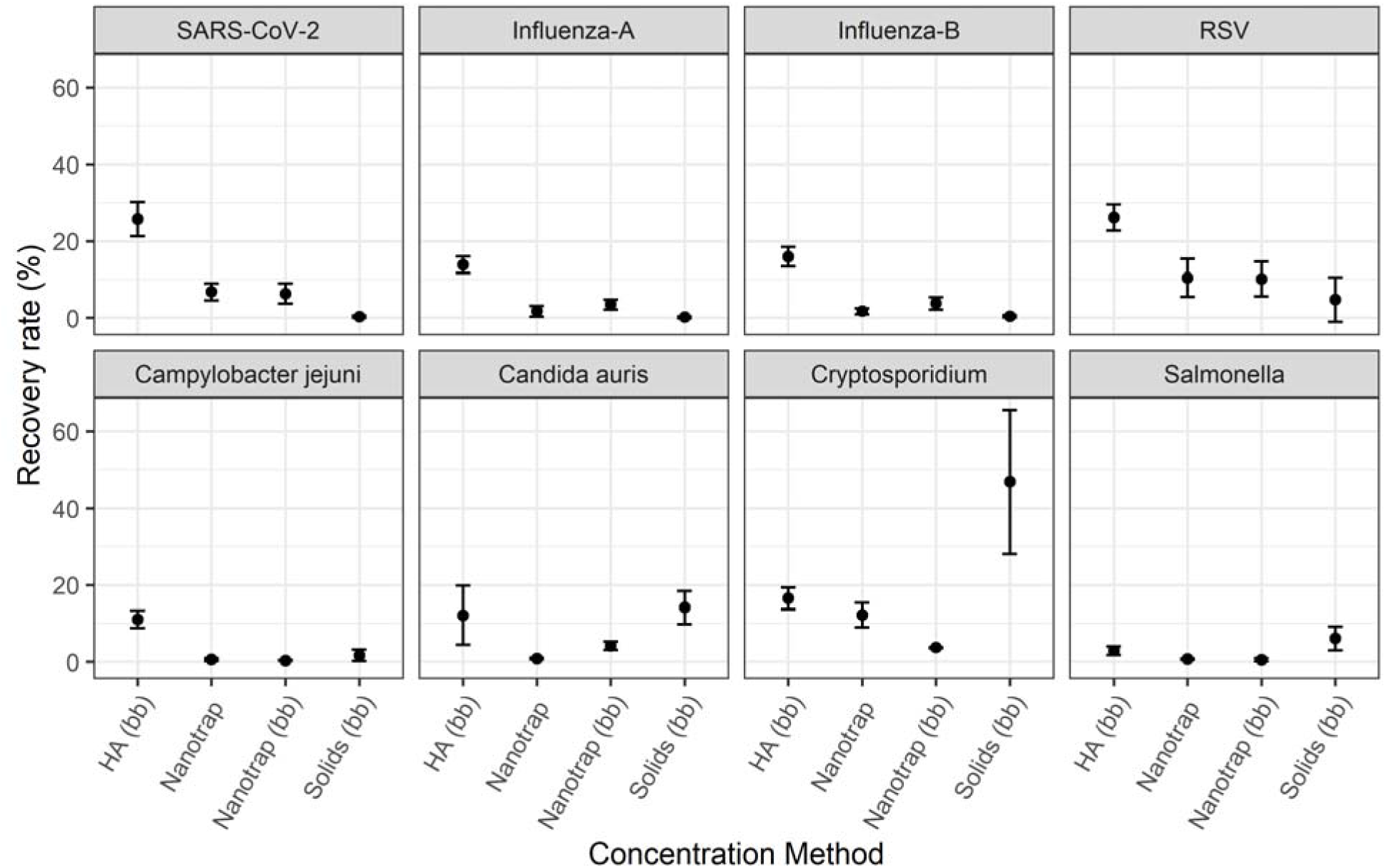
Recovery rates of target pathogens in wastewater across different concentration methods. Wastewater concentration methods included HA filtration, Nanotrap method (with or without bead-beating), and solids method. The recovery rates were calculated based on the measured pathogen concentration in wastewater and the amount of target pathogen spiked into the wastewater samples (SI.2.1).

For *Candida auris*, *Cryptosporidium*, and *Salmonella*, the highest recovery rates were associated with the solids method. For *Cryptosporidium*, we observed a recovery rate of 46.8 ± 18.7, the highest of all the targets and concentration methods in this study. The recovery rates were 14.1 ± 4.3% and 6.1 ± 3.1% for *Candida auris* and *Salmonella*, respectively, using the solids method. The highest recovery rate of *Campylobacter* was observed when the wastewater sample was processed using HA filtration, with a recovery rate of 11.1 ± 2.3%. For *Campylobacter*, the recovery rate decreased to 1.7 ± 1.5% when the solids method was used for wastewater concentration.

Our results indicate that both the choice of concentration method and the specific microbial target impact the inhibition and recovery during wastewater quantification. Inhibition was most significant when the samples were concentrated using the solids or Nanotrap methods. In addition, inhibition was greatest for the targets SARS-CoV-2, influenza A, influenza B, astrovirus, norovirus GI, and GII. This makes sense as inhibitors are more likely to be associated with and/or absorbed to the solids in wastewater samples, which are concentrated in the solids method and not removed in the Nanotrap method. Inhibitors can reduce the efficiency of both RT and PCR, and RT inhibition would disproportionally impact the quantification of RNA viruses.

Previous studies also reported differences in the recovery of microbial targets across different concentration methods. For example, Ahmed et al. (2020) reported the recovery rates of murine hepatitis virus (MHV), a surrogate of human coronavirus, ranging from 26.7 – 65.7% ^18^ . Liu et al. (2023) reported recovery rates of 24.1% and 21.5% for SARS-CoV-2 detection using HA filtration and Nanotrap method, respectively ^23^. Another study reported *Cryptosporidium* recovery rates using a centrifugation workflow between 7.5 – 15.8% when up to 100 mL of wastewater sample was processed ^20^. Again, differences in reported recovery rates across studies suggest that the results are driven by several factors, including wastewater composition, choice of concentration method, and target pathogen. The substantial variability in reported recovery rates across studies also underscores the need for normalization methods to improve their comparability, as noted by several other studies ^30–32^ .

Limitations of this study include the use of a single wastewater matrix. Wastewater composition can impact quantification of microorganisms as wastewaters can vary widely in the types and concentrations of different inhibitors that impact RT and ddPCR ^17,33^. Further, the removal of inhibitors is also impacted by the selection of concentration method. In addition, the partitioning of microorganisms in the solid and liquid fractions may differ in different wastewater matrices ^34^. Another limitation of this study is that the same nucleic acid extraction method was used for all wastewater concentration methods. However, certain concentration methods perform better when paired with specific nucleic acid extraction kits ^33,35–37^ .

### Cost analysis and application considerations for different wastewater concentration methods

To better understand the potential practical applications of each concentration method and to aid decision-makers optimize their wastewater processing workflows, we conducted a cost analysis for the wastewater concentration methods evaluated in this study (Table 2). Direct extraction had the lowest capital and operational costs among all the concentration methods ($2,613 capital cost and $5.86 operational costs per wastewater sample). The Nanotrap method that included the KingFisher system for automation resulted in the highest costs ($92,972 capital cost and $52.93 operational costs per wastewater sample). Adding a bead-beating step to the direct extraction or Nanotrap method increased the capital cost by $7,508 and increased operational costs by $2.98 per sample. Using the HA filtration method and the solids method resulted in similar costs. Capital costs were $26,496 and $24,585 for the HA filtration and solids method, respectively, and operational costs were $17.63 (HA filtration) and $16.54 (solids method) per sample.

**Table 2.** Summary of capital and operational (consumable and labor) costs of several wastewater concentration methods. The costs of concentration and bead-beating are presented separately for each method. The capital cost represents the sum of the equipment costs. The operational cost represents the operational costs for wastewater concentration per sample, assuming each sample is processed in duplicate. The operational cost is the sum of consumable and labor costs. We underlined the costs associated with the concentration methods evaluated in this study. For direct extraction and the Nanotrap method, we evaluated the cost with and without the bead-beating step. For HA filtration and the solids method, we only evaluated the costs with the bead-beating step included.

| Concentration method | | Capital cost (\$) | Consumable cost (\$) | Labor cost (\$) | Operational cost (\$) |
| --- | --- | --- | --- | --- | --- |
| **Direct extraction** | <u>Concentration</u> | <u>2,612.48</u> | *0.79* | *5.07* | <u>5.86</u> |
|  | Bead-beating | 7,507.76 | *0.73* | *2.25* | 2.98 |
|  | <u>Sum</u> | <u>10,120.24</u> | *1.52* | *7.32* | <u>8.84</u> |
| **HA filtration** | Concentration | 18,987.76 | *4.51* | *10.14* | 14.65 |
|  | Bead-beating | 7,507.76 | *0.73* | *2.25* | 2.98 |
|  | <u>Sum</u> | <u>26,495.52</u> | *5.24* | *12.39* | <u>17.63</u> |
| **Solids** | Concentration | 17,076.80 | *3.42* | *10.14* | 13.56 |
|  | Bead-beating | 7,507.76 | *0.73* | *2.25* | 2.98 |
|  | <u>Sum</u> | <u>24,584.56</u> | *4.15* | *12.39* | <u>16.54</u> |
| **HA filtration + solids** | Concentration | 18,987.76 | *6.09* | *10.70* | 16.80 |
|  | Bead-beating | 7,507.76 | *1.46* | *4.51* | 5.96 |
|  | <u>Sum</u> | <u>26,495.52</u> | *7.55* | *15.21* | <u>22.76</u> |
| **Nanotrap-kingfisher** | <u>Concentration</u> | <u>92,971.84</u> | *47.30* | *5.63* | <u>52.93</u> |
|  | Bead-beating | 7,507.76 | *0.73* | *2.25* | 2.98 |
|  | <u>Sum</u> | <u>100,479.60</u> | *48.03* | *7.89* | <u>55.91</u> |

Notably, processing wastewater samples using both HA filtration and solids methods does not substantially increase the total capital cost, and increases operational costs by $5.13 per sample as compared to HA filtration alone, as these two methods share the majority of processing steps. Considering the differences in performance across the diverse target microorganisms between HA filtration and solids methods, using a combination of the two methods, where HA filtration of the supernatant is used for viral targets and the solids are used for other microbial targets, could be a sensitive and cost-effective approach for monitoring numerous targets in each sample. The primary disadvantages associated with the HA filtration and solids methods are that they are relatively labor intensive and require substantial laboratory space for the instruments, which may be unavailable in certain laboratories. In contrast, while the Nanotrap method automates much of the process with a KingFisher system that is relatively compact and thus amenable to high throughput workflows that can process large numbers of wastewater samples, it comes at a significant cost, driven by both the capital cost of the Kingfisher and the operational costs associated with the Nanotrap beads. In summary, the selection and application of wastewater concentration methods requires balancing trade-offs in detection sensitivity, testing capacity and throughput, labor costs, and available resources.

## Conclusion

In this study, we evaluated the performance of widely used concentration methods employed for wastewater monitoring for multiple pathogens. We performed a head-to-head comparison of six different concentration methods: direct extraction with bead beating, direct extraction without bead beating, HA filtration method, solids method, Nanotrap method with bead beating, and Nanotrap method without bead beating. We compared the quantification of fourteen diverse pathogens, including respiratory viruses, pathogenic microorganisms, and enteric viruses. Positive detections were observed for all the targeted pathogens when the wastewater sample was concentrated using HA filtration, solids, or Nanotrap methods. Our results demonstrate that the choice of concentration methods significantly impacts the sensitivity of the workflow used in wastewater monitoring, driving both the theoretical LOD and the observed concentrations in wastewater samples. Our results showed that HA filtration performed best for the measurement of *Campylobacter jejuni*, adenovirus, and all target respiratory viruses. The solids method was the most sensitive for *Candida auris*, *Cryptosporidium*, and *Salmonella*. The HA filtration and Nanotrap methods performed similarly for the detection of astrovirus, sapovirus, norovirus GI, and norovirus GII in wastewater. The Nanotrap method provided the most sensitive detection for rotavirus. We do not recommend adding a bead-beating step to the direct extraction or the Nanotrap methods for the pathogens targeted in this study. Inhibition was most substantial using the solids and Nanotrap methods, and the RNA viruses were the most sensitive to inhibitors. Based on the multiplexing ability of the assays, and the performance of the different concentration methods, including the measured concentrations, impact of inhibition, and economic costs, we recommend concentrating using a combination of the HA filtration and solids methods. Extracts from the HA filtration method can be used for monitoring respiratory and enteric viruses, and the extracts from the solids method used for detecting the other target microorganisms. We realize that many laboratories prefer a more automated process and have invested in magnetic bead-based concentration methods. Our results show that the Nanotrap method is also very effective for wastewater monitoring when a high-throughput system is needed. However, using the Nanotrap method will result in less sensitivity for certain pathogens and higher economic costs. Innovation is needed to develop automated filtration-based concentration systems that reduce hands-on labor time and competitive technologies to the Nanotrap beads that drive costs down. The findings of this study can inform the development and optimization of wastewater surveillance systems for monitoring different pathogens, providing valuable data to support public health responses for a wide range of diseases.

## Data Availability

All data produced in the present study are available upon reasonable request to the authors.

## Acknowledgments

This research was supported by funds from the Centers for Disease Control and Prevention (NI50CK000557) and the Houston Health Department. Todd J. Treangen and Michael X. Wang were supported in part by National Science Foundation (NSF) award IIS-2239114. We thank Kaavya Domakonda and Rebecca Schneider at the Houston Health Department and Houston Water for their assistance in sample collection. We also thank Dolores Sanchez Gonzalez and Michael Secreto for collecting and sending wastewater samples and for an insightful conversation. We thank Lauren Bauhs, Robert Campos, and Kyle Palmer for their help with wastewater processing. We also thank Kiara Reyes Gamas for her contributions to the graphical abstract and Figure 1. The graphical abstract was created with Biorender.com.

## References

(1) Barber, C.; Crank, K.; Papp, K.; Innes, G. K.; Schmitz, B. W.; Chavez, J.; Rossi, A.; Gerrity, D. Community-Scale Wastewater Surveillance of Candida Auris during an Ongoing Outbreak in Southern Nevada. Environ. Sci. Technol. 2023, 57 (4), 1755–1763. 10.1021/acs.est.2c07763.

(2) Boehm, A. B.; Wolfe, M. K.; White, B. J.; Hughes, B.; Duong, D.; Bidwell, A. More than a Tripledemic: Influenza A Virus, Respiratory Syncytial Virus, SARS-CoV-2, and Human Metapneumovirus in Wastewater during Winter 2022–2023. Environ. Sci. Technol. Lett. 2023, 10 (8), 622–627. 10.1021/acs.estlett.3c00385.

(3) Boehm, A. B.; Wolfe, M. K.; White, B. J.; Hughes, B.; Duong, D. Two Years of Longitudinal Measurements of Human Adenovirus Group F, Norovirus GI and GII, Rotavirus, Enterovirus, Enterovirus D68, Hepatitis A Virus, Candida Auris, and West Nile Virus Nucleic-Acids in Wastewater Solids: A Retrospective Study at Two Wastewater Treatment Plants; preprint; Epidemiology, 2023. 10.1101/2023.08.22.23294424.

(4) de Melo, T.; Islam, G.; Simmons, D. B. D.; Desaulniers, J.-P.; Kirkwood, A. E. An Alternative Method for Monitoring and Interpreting Influenza A in Communities Using Wastewater Surveillance. Front. Public Health 2023, 11, 1141136. 10.3389/fpubh.2023.1141136.

(5) Dumke, R.; Geissler, M.; Skupin, A.; Helm, B.; Mayer, R.; Schubert, S.; Oertel, R.; Renner, B.; Dalpke, A. H. Simultaneous Detection of SARS-CoV-2 and Influenza Virus in Wastewater of Two Cities in Southeastern Germany, January to May 2022. Int. J. Environ. Res. Public. Health 2022, 19 (20), 13374. 10.3390/ijerph192013374.

(6) Hayes, E. K.; Gouthro, M. T.; LeBlanc, J. J.; Gagnon, G. A. Simultaneous Detection of SARS-CoV-2, Influenza A, Respiratory Syncytial Virus, and Measles in Wastewater by Multiplex RT-qPCR. Sci. Total Environ. 2023, 889, 164261. 10.1016/j.scitotenv.2023.164261.

(7) Huang, Y.; Zhou, N.; Zhang, S.; Yi, Y.; Han, Y.; Liu, M.; Han, Y.; Shi, N.; Yang, L.; Wang, Q.; Cui, T.; Jin, H. Norovirus Detection in Wastewater and Its Correlation with Human Gastroenteritis: A Systematic Review and Meta-Analysis. Environ. Sci. Pollut. Res. Int. 2022, 29 (16), 22829–22842. 10.1007/s11356-021-18202-x.

(8) Hughes, B.; Duong, D.; White, B. J.; Wigginton, K. R.; Chan, E. M. G.; Wolfe, M. K.; Boehm, A. B. Respiratory Syncytial Virus (RSV) RNA in Wastewater Settled Solids Reflects RSV Clinical Positivity Rates. Environ. Sci. Technol. Lett. 2022, 9 (2), 173–178. 10.1021/acs.estlett.1c00963.

(9) Koureas, M.; Mellou, K.; Vontas, A.; Kyritsi, M.; Panagoulias, I.; Koutsolioutsou, A.; Mouchtouri, V. A.; Speletas, M.; Paraskevis, D.; Hadjichristodoulou, C. Wastewater Levels of Respiratory Syncytial Virus Associated with Influenza-like Illness Rates in Children—A Case Study in Larissa, Greece (October 2022–January 2023). Int. J. Environ. Res. Public. Health 2023, 20 (6), 5219. 10.3390/ijerph20065219.

(10) Lee, A. J.; Carson, S.; Reyne, M. I.; Marshall, A.; Moody, D.; Allen, D. M.; Allingham, P.; Levickas, A.; Fitzgerald, A.; Bell, S. H.; Lock, J.; Coey, J. D.; McSparron, C.; Nejad, B. F.; Courtney, D. G.; Einarsson, G. G.; McKenna, J. P.; Fairley, D. J.; Curran, T.; McKinley, J. M.; Gilpin, D. F.; Lemon, K.; McGrath, J. W.; Bamford, C. G. G. ‘One Health’ Genomic Surveillance of Avian and Human Influenza A Viruses Through Environmental Wastewater Monitoring. medRxiv August 13, 2023, p 2023.08.08.23293833. 10.1101/2023.08.08.23293833.

(11) Markt, R.; Stillebacher, F.; Nägele, F.; Kammerer, A.; Peer, N.; Payr, M.; Scheffknecht, C.; Dria, S.; Draxl-Weiskopf, S.; Mayr, M.; Rauch, W.; Kreuzinger, N.; Rainer, L.; Bachner, F.; Zuba, M.; Ostermann, H.; Lackner, N.; Insam, H.; Wagner, A. O. Expanding the Pathogen Panel in Wastewater Epidemiology to Influenza and Norovirus. Viruses 2023, 15 (2), 263. 10.3390/v15020263.

(12) Mercier, E.; D’Aoust, P. M.; Thakali, O.; Hegazy, N.; Jia, J.-J.; Zhang, Z.; Eid, W.; Plaza-Diaz, J.; Kabir, M. P.; Fang, W.; Cowan, A.; Stephenson, S. E.; Pisharody, L.; MacKenzie, A. E.; Graber, T. E.; Wan, S.; Delatolla, R. Municipal and Neighbourhood Level Wastewater Surveillance and Subtyping of an Influenza Virus Outbreak. Sci. Rep. 2022, 12 (1), 15777. 10.1038/s41598-022-20076-z.

(13) Wolfe, M. K.; Duong, D.; Bakker, K. M.; Ammerman, M.; Mortenson, L.; Hughes, B.; Arts, P.; Lauring, A. S.; Fitzsimmons, W. J.; Bendall, E.; Hwang, C. E.; Martin, E. T.; White, B. J.; Boehm, A. B.; Wigginton, K. R. Wastewater-Based Detection of Two Influenza Outbreaks. Environ. Sci. Technol. Lett. 2022. 10.1021/acs.estlett.2c00350.

(14) Wolken, M.; Wang, M.; Schedler, J.; Campos, R. H.; Ensor, K.; Hopkins, L.; Treangen, T.; Stadler, L. B. PreK-12 School and Citywide Wastewater Monitoring of the Enteric Viruses Astrovirus, Rotavirus, and Sapovirus. Sci. Total Environ. 2024, 931, 172683. 10.1016/j.scitotenv.2024.172683.

(15) Wolken, M.; Sun, T.; McCall, C.; Schneider, R.; Caton, K.; Hundley, C.; Hopkins, L.; Ensor, K.; Domakonda, K.; Kalvapalle, P.; Persse, D.; Williams, S.; Stadler, L. B. Wastewater Surveillance of SARS-CoV-2 and Influenza in preK-12 Schools Shows School, Community, and Citywide Infections. Water Res. 2023, 231, 119648. 10.1016/j.watres.2023.119648.

(16) Zafeiriadou, A.; Kaltsis, L.; Kostakis, M.; Kapes, V.; Thomaidis, N. S.; Markou, A. Wastewater Surveillance of the Most Common Circulating Respiratory Viruses in Athens: The Impact of COVID-19 on Their Seasonality. Sci. Total Environ. 2023, 900, 166136. 10.1016/j.scitotenv.2023.166136.

(17) Ahmed, W.; Simpson, S. L.; Bertsch, P. M.; Bibby, K.; Bivins, A.; Blackall, L. L.; Bofill-Mas, S.; Bosch, A.; Brandão, J.; Choi, P. M.; Ciesielski, M.; Donner, E.; D’Souza, N.; Farnleitner, A. H.; Gerrity, D.; Gonzalez, R.; Griffith, J. F.; Gyawali, P.; Haas, C. N.; Hamilton, K. A.; Hapuarachchi, H. C.; Harwood, V. J.; Haque, R.; Jackson, G.; Khan, S. J.; Khan, W.; Kitajima, M.; Korajkic, A.; La Rosa, G.; Layton, B. A.; Lipp, E.; McLellan, S. L.; McMinn, B.; Medema, G.; Metcalfe, S.; Meijer, W. G.; Mueller, J. F.; Murphy, H.; Naughton, C. C.; Noble, R. T.; Payyappat, S.; Petterson, S.; Pitkänen, T.; Rajal, V. B.; Reyneke, B.; Roman, F. A.; Rose, J. B.; Rusiñol, M.; Sadowsky, M. J.; Sala-Comorera, L.; Setoh, Y. X.; Sherchan, S. P.; Sirikanchana, K.; Smith, W.; Steele, J. A.; Sabburg, R.; Symonds, E. M.; Thai, P.; Thomas, K. V.; Tynan, J.; Toze, S.; Thompson, J.; Whiteley, A. S.; Wong, J. C. C.; Sano, D.; Wuertz, S.; Xagoraraki, I.; Zhang, Q.; Zimmer-Faust, A. G.; Shanks, O. C. Minimizing Errors in RT-PCR Detection and Quantification of SARS-CoV-2 RNA for Wastewater Surveillance. Sci. Total Environ. 2022, 805, 149877. 10.1016/j.scitotenv.2021.149877.

(18) Ahmed, W.; Bertsch, P. M.; Bivins, A.; Bibby, K.; Farkas, K.; Gathercole, A.; Haramoto, E.; Gyawali, P.; Korajkic, A.; McMinn, B. R.; Mueller, J. F.; Simpson, S. L.; Smith, W. J. M.; Symonds, E. M.; Thomas, K. V.; Verhagen, R.; Kitajima, M. Comparison of Virus Concentration Methods for the RT-qPCR-Based Recovery of Murine Hepatitis Virus, a Surrogate for SARS-CoV-2 from Untreated Wastewater. Sci. Total Environ. 2020, 739, 139960. 10.1016/j.scitotenv.2020.139960.

(19) Ciannella, S.; González-Fernández, C.; Gomez-Pastora, J. Recent Progress on Wastewater-Based Epidemiology for COVID-19 Surveillance: A Systematic Review of Analytical Procedures and Epidemiological Modeling. Sci. Total Environ. 2023, 878, 162953. 10.1016/j.scitotenv.2023.162953.

(20) Hachimi, O.; Falender, R.; Davis, G.; Wafula, R. V.; Sutton, M.; Bancroft, J.; Cieslak, P.; Kelly, C.; Kaya, D.; Radniecki, T. Evaluation of Molecular-Based Methods for the Detection and Quantification of *Cryptosporidium* Spp. in Wastewater. Sci. Total Environ. 2024, 947, 174219. 10.1016/j.scitotenv.2024.174219.

(21) Laturner, Z. W.; Zong, D. M.; Kalvapalle, P.; Gamas, K. R.; Terwilliger, A.; Crosby, T.; Ali, P.; Avadhanula, V.; Santos, H. H.; Weesner, K.; Hopkins, L.; Piedra, P. A.; Maresso, A. W.; Stadler, L. B. Evaluating Recovery, Cost, and Throughput of Different Concentration Methods for SARS-CoV-2 Wastewater-Based Epidemiology. Water Res. 2021, 197, 117043. 10.1016/j.watres.2021.117043.

(22) Ahmed, W.; Bivins, A.; Korajkic, A.; Metcalfe, S.; Smith, W. J. M.; Simpson, S. L. Comparative Analysis of Adsorption-Extraction (AE) and Nanotrap® Magnetic Virus Particles (NMVP) Workflows for the Recovery of Endogenous Enveloped and Non-Enveloped Viruses in Wastewater. Sci. Total Environ. 2023, 859, 160072. 10.1016/j.scitotenv.2022.160072.

(23) Liu, P.; Guo, L.; Cavallo, M.; Cantrell, C.; Hilton, S. P.; Nguyen, A.; Long, A.; Dunbar, J.; Barbero, R.; Barclay, R.; Sablon, O.; Wolfe, M.; Lepene, B.; Moe, C. Comparison of Nanotrap® Microbiome A Particles, Membrane Filtration, and Skim Milk Workflows for SARS-CoV-2 Concentration in Wastewater. Front. Microbiol. 2023, 14. 10.3389/fmicb.2023.1215311.

(24) Zheng, X.; Zhao, K.; Xu, X.; Deng, Y.; Leung, K.; Wu, J. T.; Leung, G. M.; Peiris, M.; Poon, L. L. M.; Zhang, T. Development and Application of Influenza Virus Wastewater Surveillance in Hong Kong. Water Res. 2023, 245, 120594. 10.1016/j.watres.2023.120594.

(25) Ahmed, W.; Bivins, A.; Simpson, S. L.; Smith, W. J. M.; Metcalfe, S.; McMinn, B.; Symonds, E. M.; Korajkic, A. Comparative Analysis of Rapid Concentration Methods for the Recovery of SARS-CoV-2 and Quantification of Human Enteric Viruses and a Sewage-Associated Marker Gene in Untreated Wastewater. Sci. Total Environ. 2021, 799, 149386. 10.1016/j.scitotenv.2021.149386.

(26) Lou, E. G.; Sapoval, N.; McCall, C.; Bauhs, L.; Carlson-Stadler, R.; Kalvapalle, P.; Lai, Y.; Palmer, K.; Penn, R.; Rich, W.; Wolken, M.; Brown, P.; Ensor, K. B.; Hopkins, L.; Treangen, T. J.; Stadler, L. B. Direct Comparison of RT-ddPCR and Targeted Amplicon Sequencing for SARS-CoV-2 Mutation Monitoring in Wastewater. Sci. Total Environ. 2022, 833, 155059. 10.1016/j.scitotenv.2022.155059.

(27) Borchardt, M. A.; Boehm, A. B.; Salit, M.; Spencer, S. K.; Wigginton, K. R.; Noble, R. T. The Environmental Microbiology Minimum Information (EMMI) Guidelines: qPCR and dPCR Quality and Reporting for Environmental Microbiology. Environ. Sci. Technol. 2021, 55 (15), 10210–10223. 10.1021/acs.est.1c01767.

(28) Cao, Y.; Griffith, J. F.; Dorevitch, S.; Weisberg, S. B. Effectiveness of qPCR Permutations, Internal Controls and Dilution as Means for Minimizing the Impact of Inhibition While Measuring Enterococcus in Environmental Waters. J. Appl. Microbiol. 2012, 113 (1), 66–75. 10.1111/j.1365-2672.2012.05305.x.

(29) Kang, T. S. Basic Principles for Developing Real-Time PCR Methods Used in Food Analysis: A Review. Trends Food Sci. Technol. 2019, 91, 574–585. 10.1016/j.tifs.2019.07.037.

(30) Endo, N.; Hisahara, A.; Kameda, Y.; Mochizuki, K.; Kitajima, M.; Yasojima, M.; Daigo, F.; Takemori, H.; Nakamura, M.; Matsuda, R.; Iwamoto, R.; Nojima, Y.; Ihara, M.; Tanaka, H. Enabling Quantitative Comparison of Wastewater Surveillance Data across Methods through Data Standardization without Method Standardization. Sci. Total Environ. 2024, 953, 176073. 10.1016/j.scitotenv.2024.176073.

(31) Islam, G.; Gedge, A.; Ibrahim, R.; de Melo, T.; Lara-Jacobo, L.; Dlugosz, T.; Kirkwood, A. E.; Simmons, D.; Desaulniers, J.-P. The Role of Catchment Population Size, Data Normalization, and Chronology of Public Health Interventions on Wastewater-Based COVID-19 Viral Trends. Sci. Total Environ. 2024, 937, 173272. 10.1016/j.scitotenv.2024.173272.

(32) Mazumder, P.; Dash, S.; Honda, R.; Sonne, C.; Kumar, M. Sewage Surveillance for SARS-CoV-2: Molecular Detection, Quantification, and Normalization Factors. Curr. Opin. Environ. Sci. Health 2022, 28, 100363. 10.1016/j.coesh.2022.100363.

(33) S. Antkiewicz, D.; H. Janssen, K.; Roguet, A.; E. Pilch, H.; B. Fahney, R.; A. Mullen, P.; N. Knuth, G.; G. Everett, D.; M. Doolittle, E.; King, K.; Wood, C.; Stanley, A.; C. Hemming, J. D.; M. Shafer, M. Wastewater-Based Protocols for SARS-CoV-2: Insights into Virus Concentration, Extraction, and Quantitation Methods from Two Years of Public Health Surveillance. Environ. Sci. Water Res. Technol. 2024, 10 (8), 1766–1784. 10.1039/D3EW00958K.

(34) Roldan-Hernandez, L.; Boehm, A. B. Adsorption of Respiratory Syncytial Virus, Rhinovirus, SARS-CoV-2, and F+ Bacteriophage MS2 RNA onto Wastewater Solids from Raw Wastewater. Environ. Sci. Technol. 2023, 57 (36), 13346–13355. 10.1021/acs.est.3c03376.

(35) Chandra, F.; Armas, F.; Kwok, G.; Chen, H.; Desmond Chua, F. J.; Leifels, M.; Amir-Hamzah, N.; Gu, X.; Wuertz, S.; Lee, W. L.; Alm, E. J.; Thompson, J. Comparing Recovery Methods for Wastewater Surveillance of Arthropod-Borne and Enveloped Viruses. ACS EST Water 2023, 3 (4), 974–983. 10.1021/acsestwater.2c00460.

(36) Kim, S.; Garcia, D.; McCormack, C.; Tham, R. X.; O’Brien, M. E.; Fuhrmeister, E. R.; Nelson, K. L.; Kantor, R. S.; Pickering, A. J. Solid Evidence and Liquid Gold: Trade-Offs of Processing Settled Solids, Whole Influent, or Centrifuged Influent for Co-Detecting Viral, Bacterial, and Eukaryotic Pathogens in Wastewater. medRxiv March 5, 2025, p 2025.03.01.25323156. 10.1101/2025.03.01.25323156.

(37) O’Brien, M.; Rundell, Z. C.; Nemec, M. D.; Langan, L. M.; Back, J. A.; Lugo, J. N. A Comparison of Four Commercially Available RNA Extraction Kits for Wastewater Surveillance of SARS-CoV-2 in a College Population. Sci. Total Environ. 2021, 801, 149595. 10.1016/j.scitotenv.2021.149595.

