## Supplementary material for "Sensitivity, throughput, and cost analysis of concentration methods for multi-target pathogen wastewater monitoring": SI

### **SI 1. Materials and Methods**

#### **SI 1.1 Assay development for *Adenovirus* 41 and *Cryptosporidium***

130 Human Adenovirus F41 genomes were downloaded from the NCBI Virus database ^1^, with genome length greater than 30k bp and release date before April 14, 2023. PCR primers and the TaqMan probe were designed on the conserved regions of the short fiber gene (NCBI accession number AYV33729.1) using Primer3Plus ^2^. Primer and probe sequences are also checked for specificity with Olivar ^3^ against the F40 serotype, as well as other species under Mastadenovirus, including Human mastadenovirus A, B, C, D, E and G, showing sufficient specificity to the F41 serotype. For *Cryptosporidium*, 4 complete, annotated genomes under *Cryptosporidium* *parvum* were downloaded from GenBank, including 1 RefSeq reference genome. PCR primers and the TaqMan probe were designed using Primer3Plus based on the consensus of all genome assemblies, targeting the Ataxin-2 related NUDIX domain protein (accession number XM_628439.1). The designed assay specifically detects *Cryptosporidium parvum* and *Cryptosporidium hominis*, which are the species of primary concern for human health ^4^. Primer and probe sequences were then checked against non-target sequences with Olivar ^3^ using the BLAST nt database ^5^.

#### **SI 1.2 Wastewater sample processing**

The pathogen standards (Table 1) were diluted and mixed before being spiked into wastewater bottle A. Wastewater samples were homogenized by inverting the bottle containing the sample several times. The concentrations of pathogen standards in the spike-in solution were processed using three different methods before being quantified using (RT-)ddPCR as described below. The pre-treatments include: 1) heat-lysis at 95ºC for 10 minutes using C1000 Touch™ Thermal Cycler (1851197, Bio-Rad); 2) direct nucleic acid extraction using Chemagic^TM^ Prime Viral DNA/RNA 300 Kit H96 (Chemagic, CMG-1433, PerkinElmer); and 3) transferring 10 µL of the standard solution to a filled bead beating tube containing 0.1 mm diameter glass beads and 1000 µL of lysis buffer, bead beating (described below), and the nucleic acid extraction using Chemagic^TM^ Prime Viral DNA/RNA 300 Kit H96 (Chemagic, CMG-1433, PerkinElmer). The quantification with each pre-treatment process was performed in triplicate.

For direct extraction, 1 mL of wastewater sample was aliquoted into 1.5 mL centrifuge tubes. The tubes were centrifuged at 17,000 g and 4°C for 5 minutes. After centrifugation, 300 µL of the supernatant was directly used for nucleic acid extraction and 300 µL of the supernatant was carefully transferred to a filled bead beating tube containing 0.1 mm diameter glass beads and 700 µL of lysis buffer from the chemagic^TM^ Prime Viral DNA/RNA 300 Kit H96 (CMG-1433, PerkinElmer) without disturbing the pellet.

For the HA filtration and solids method, the wastewater sample was aliquoted into 50 mL centrifuge tubes. Tubes containing wastewater were centrifuged at 4,100 g and 4ºC for 20 minutes to separate liquid and solid fractions. The wastewater liquid fraction concentration was described previously by Laturner et al. (2021) and Lou et al. (2022) and summarized as follows ^6,7^. After centrifugation, the supernatant was carefully poured into an MF 3, 300ml Magnetic Filter Holder with lid kit (20030001, Sterlitech) attached to the Multi-Vac 600-MS Manifold (180600-01, Sterlitech) and Rocker 800 Oil Free Laboratory Vacuum Pump (167800, Sterlitech) system without disturbing the pellet. The Electronegative Microbiological Analysis Membrane HA Filter (HAWG047S6, Millipore Sigma) was placed into the manifold system prior to the sample addition. One mL of 1.25 M MgCl_2_·H_2_O (M0250, Sigma Aldrich) solution was added to the sample in the filter cup and then gently swirled with the pipette tip to mix and allowed to stand for 5 minutes before the vacuum pump was turned on. After all the supernatant passed through the filter, the filter was folded and transferred into a filled bead beating tube containing 0.1 mm diameter glass beads and 1 mL of lysis buffer from the chemagic^TM^ Prime Viral DNA/RNA 300 Kit H96 (CMG-1433, PerkinElmer), which was used for nucleic acid extraction. For the wastewater solid fraction, the pellet that remained at the bottom of the tube was resuspended using 1 mL of lysis buffer from the Chemagic^TM^ Prime Viral DNA/RNA 300 Kit H96 (CMG-1433, PerkinElmer) and transferred into a filled bead beating tube containing 0.1 mm diameter glass beads.

The Nanotrap method was conducted following the manufacturer’s manual and summarized as follows: 10 mL wastewater sample was aliquoted into 15 mL concentration tubes. 100 µL of Nanotrap Enhancement Reagent 3 (10113, Ceres Nanosciences), 150 µL of Nanotrap Microbiome A Particles (44202, Ceres Nanosciences), and 150 µL of Nanotrap Microbiome B Particles (65202, Ceres Nanosciences) were added into the sample, mixed, and incubated at room temperature for 10 minutes. A DynaMag-15 magnetic rack (12301D, Thermo Fisher Scientific) was used to separate the Nanotrap particles, and the supernatant was carefully discarded without disturbing the Nanotrap particle pellet. 1 mL Nanotrap Buffer 2 (9999, Ceres Nanosciences) was used to resuspend the Nanotrap particle pellet before being transferred to 2 mL microcentrifuge tubes. The tubes were placed on a DynaMag-2 magnetic rack (12321D, Thermo Fisher Scientific) to separate the Nanotrap particles. The supernatant was discarded before 600 µL of lysis buffer from the Chemagic^TM^ Prime Viral DNA/RNA 300 Kit H96 (CMG-1433, PerkinElmer) was added to mix with the Nanotrap particles. The tubes were then incubated at 95ºC for 10 minutes and placed on the DynaMag-2 magnetic rack (12321D, Thermo Fisher Scientific) to separate the Nanotrap particles. 300 µL of the supernatant was directly used for nucleic acid extraction and 300 µL of the supernatant was carefully transferred to a filled bead beating tube containing 0.1 mm diameter glass beads and 700 µL of lysis buffer from the Chemagic^TM^ Prime Viral DNA/RNA 300 Kit H96 (CMG-1433, PerkinElmer).

For the bead-beating step, the bead-beating tubes were beaten for 1 minute at 3,500 oscillations/m two times using the Mini-Beadbeater 24 (112011, BioSpec) with a 2-minute break, where the tubes were placed on ice. After bead beating, the tubes were centrifuged at 17,000 g and 4°C for 5 minutes. 300 µL of lysate was used as input to nucleic acid extraction using Chemagic^TM^ Prime Viral DNA/RNA 300 Kit H96 (Chemagic, CMG-1433, PerkinElmer), following the manufacturer’s protocol, and the extracted nucleic acids were eluted in 50 µL of sterile, nuclease-free water. The extracts were stored at 4°C for no more than 2 hours before quantification.

The concentrations of target RNA were quantified using one-step RT-ddPCR Advanced Kit for Probes (1864021, Bio-Rad). The concentrations of target DNA were quantified using ddPCR Multiplex Supermix (12005910, Bio-Rad). Droplet generation was performed using an Automated Droplet Generator (1864101, Bio-Rad) and RT-ddPCR was completed using a C1000 ThermalCycler (1851197, Bio-Rad) and QX600 AutoDG Droplet Digital PCR System (12013328, Bio-Rad). Results were analyzed using QuantaSoft v1.7.4 software.

#### **SI 1.3 Concentration factor calculations and equivalent volume for each method**

**Table SI.1:** Concentration factor for direct extraction without bead beating

|  | **Volume** |  | **Concentration factor** |
| --- | --- | --- | --- |
| Sample volume: | 300 | µL |  |
| Elution volume: | 50 | µL | 6 |
|  |  | **Total:** | 6 |

**Table SI.2:** Concentration factor for direct extraction with bead beating

|  | **Volume** |  | **Concentration factor** |
| --- | --- | --- | --- |
| Sample volume: | 300 | µL |  |
| Lysis buffer added | 1000 | µL | 0.3 |
| Added to Chemagic | 300 | µL |  |
| Elution volume: | 50 | ul | 6 |
|  |  | **Total:** | 1.8 |

**Table SI.3:** Concentration factor for HA filtration and solids method

|  | **Volume** |  | **Concentration factor** |
| --- | --- | --- | --- |
| Sample volume: | 50 | mL |  |
| Lysis buffer added | 1000 | µL | 50 |
| Added to Chemagic | 300 | µL |  |
| Elution volume: | 50 | ul | 6 |
|  |  | **Total:** | 300 |

**Table SI.4:** Concentration factor for Nanotrap method without bead beating

|  | **Volume** |  | **Concentration factor** |
| --- | --- | --- | --- |
| Sample volume: | 10 | mL |  |
| Lysis buffer added | 600 | µL | 16.67 |
| Added to Chemagic | 300 | µL |  |
| Elution volume: | 50 | ul | 6 |
|  |  | **Total:** | 100 |

**Table SI.5:** Concentration factor for Nanotrap method with bead beating

|  | **Volume** |  | **Concentration factor** |
| --- | --- | --- | --- |
| Sample volume: | 10 | mL |  |
| Lysis buffer added | 600 | µL | 16.67 |
| Volume for bead beating | 300 | µL |  |
| Lysis buffer added for bead beating | 1000 | µL | 0.3 |
| Added to Chemagic | 300 | µL |  |
| Elution volume: | 50 | ul | 6 |
|  |  | **Total:** | 30 |

#### **SI 1.4 (RT-)ddPCR assays and thermal cycling conditions**

**Table SI.6:** ddPCR assays and positive standards used for quantification

| **Virus/Target^*^** | **Assay name** | **Sequence (5’-3’)** |
| --- | --- | --- |
| SARS-CoV-2 ^8^ | Forward primer | TTACAAACATTGGCCGCAAA |
|  | Reverse primer | GCGCGACATTCCGAAGAA |
|  | Probe | Cy55/ACAATTTGCCCCCAGCGCTTCAG/3IAbRQSp |
|  | Amplicon length | 67 |
|  | Gblock sequence | CATACAATGTAACACAAGCTTTCGGCAGACGTGGTCCAGAACAAACCCAAGGAAATTTTGGGGACCAGGAACTAATCAGACAAGGAACTGATTACAAACATTGGCCGCAAATTGCACAATTTGCCCCCAGCGCTTCAGCGTTCTTCGGAATGTCGCGCATTGGCATGGAAGTCACACCTTCGGGAACGTGGTTGACCTACACAGGTGCCATCAAATTGGATGACAAAGATCCAAATTTCAAAGATCAAGTC |
| Influenza A ^9^ | Forward primer | CTTCTAACCGAGGTCGAAACGTA |
|  | Reverse primer | GGTGACAGGATTGGTCTTGTCTTTA |
|  | Probe | SUN/TCAGGCCCC/ZEN/CTCAAAGCCGAG/3IABkFQ |
|  | Amplicon length | 155 |
|  | Gblock sequence | AGGGTCTCGCGACATGAGTCTTCTAACCGAGGTCGAAACGTACGTTCTCTCTATCGTCCCGTCAGGCCCCCTCAAAGCCGAGATCGCGCAGAGACTTGAAGATGTGTTTGCAGGGAAGAACACCGATCTTGAGGCACTCATGGAATGGCTAAAGACAAGACCAATCCTGTCACCTCTGACTAAGGGGATTTTAGGATTTGTGTTCACGCTCACCGTGCCCAGTGAGCGAGGACTGCAGCGTAGACGCTTTGTCCAAAATGCCCTTAATGGGAATGGGGATCCAAACAACATGGACAGAGCGGTCAAACTGTACATGGCAGAGACCTA |
| Influenza B ^10^ | Forward primer | AAATACGGTGGATTAAACAAAAGCAA |
|  | Reverse primer | CCAGCAATAGCTCCGAAGAAA |
|  | Probe | ROXN/CACCCATATTGGGCAATTTCCTATGGC/3IAbRQSp |
|  | Amplicon length | 170 |
|  | Gblock sequence | AGGGTCTCGCGACGTAATAAAAGGGTCCTTGCCTTTAATTGGTGAAGCAGATTGCCTCCATGAAAAATACGGTGGATTAAACAAAAGCAAGCCTTACTACACAGGAGAACATGCAAAAGCCATAGGAAATTGCCCAATATGGGTGAAAACACCCTTGAAGCTGGCCAATGGAACCAAATATAGACCGCCTGCAAAACTATTAAAGGAAAGGGGTTTCTTTGGAGCTATTGCTGGTTTCTTGGAAGGAGGATGGGAAGGAATGATTGCAGGTTGGCACGGATACACATCTCATGGAGCACATGGAGTGGCAGTGGCTGGCAGAGACCTA |
| RSV ^11^ | Forward primer | CTCCAGAATAYAGGCATGAYTCTCC |
|  | Reverse primer | GCYCTYCTAATYACWGCTGTAAGAC |
|  | Probe | FAM/TAACCAAAT/ZEN/TAGCAGCAGGAGATAGATCAG/3IABkFQ |
|  | Amplicon length | 121 |
|  | Gblock sequence | CTAGAAAATCCTACAAAAAAATGCTAAAAGAAATGGGAGAGGTAGCTCCAGAATACAGGCATGACTCTCCTGATTGTGGGATGATAATATTATGTATAGCGGCATTAGTAATAACCAAATTAGCAGCAGGAGATAGATCAGGTCTTACAGCTGTGATTAGGAGGGCTAATAATGTCCTAAAAAATGAAATGAAACGTTATAAAGGCTTACTACCCAAGGATATAGCCAACAGCTTCTATGAAGTGTTTGAAAAATATCCTCACTTTATAGATGTTTTTGTTCATTTTGGTATAGCACAATCTTCTA |
| *Cryptosporidium* | Forward primer | CCAAAACTTTCTTTGCGAGT |
|  | Reverse primer | TGCTCTTCAGGAGATGAAAC |
|  | Probe | FAM/TTGGACAAT/ZEN/TTGTTGCTGAAGATTGTCC/3IABkFQ |
|  | Amplicon length | 86 |
| *Candida auris* ^12^ | Forward Primer | CAGACGTGAATCATCGAATCT |
|  | Reverse Primer | TTTCGTGCAAGCTGTAATTT |
|  | Probe | SUN/AATCTTCGC/ZEN/GGTGGCGTTGCATTCA/3IABkFQ |
|  | Amplicon length | 135 |
| *Salmonella enterica* ^13^ | Forward primer | AGCGTACTGGAAAGGGAAAG |
|  | Reverse primer | ATACCGCCAATAAAGTTCACAAAG |
|  | Probe | Cy5/CGTCACCTT/TAO/TGATAAACTTCATCGCA/3IAbRQSp |
|  | Amplicon length | 116 |
| *Campylobacter jejuni* ^14^ | Forward primer | TGCACCAGTGACTATGAATAACGA |
|  | Reverse primer | TCCAAAATCCTCACTTGCCATT |
|  | Probe | Cy55/TTGCAACCTCACTAGCAAAATCCACAGCT/3IAbRQSp |
|  | Amplicon length | 124 |
| *Cryptosporidium* + *Candida auris* + *Salmonella enterica* + *Campylobacter jejuni* | Gblock sequence | TTTCGTGCAAGCTGTAATTTTGTGAATGCAACGCCACCGCGAAGATTGGTGAGAAGACATCACGCTCAAACAGGCATGCCTTGGGGAATACCCCAAGGCGCAATGTGCGTTCAAAGATTCGATGATTCACGTCTGCAAGTTTGCACCAGTGACTATGAATAACGATGAAGCTGTGGATTTTGCTAGTGAGGTTGCAAAAGAATTATTTGGCGAAAAAAATTGTGAATTTAATCATCGTCCTTTAATGGCAAGTGAGGATTTTGGATTAGCGTACTGGAAAGGGAAAGTCAGCTTTACGGTTCCTTTGACGGTGCGATGAAGTTTATCAAAGGTGACGCCATTGCCGGTATCATTATCATCTTTGTGAACTTTATTGGCGGTATTAAGAAATTTGTCATGCTCTTCAGGAGATGAAACGAAATGTCGTAAAATTGGACAATCTTCAGCAACAAATTGTCCAAAGACTCGCAAAGAAAGTTTTGGTAGAACATGTGA |
| Sapovirus ^15^ | Forward primer 1 | CCATCCAATCAAATGTCCCTG |
|  | Forward primer 2 | CAATCCAATCCAATGTCCCTG |
|  | Reverse primer 1 | CTGTCATTCCAAGCAAAAGTAC |
|  | Reverse primer 2 | CTGTCGTTCCAAGCAAAAGTAC |
|  | Probe | FAM/ATACGCAAC/ZEN/TGCTTTGCAGTCTKTC/3IABkFQ |
|  | Amplicon length | 73 |
|  | Gblock sequence | CCGATGGTCGTTGACCCGCCTGGCACAACAGGTCCGACCACATCCCACGTTGTTGTTGCTAATCCGGAGCAACCCAATGGGGCCGCACAGCGCCTGGAGTTGGCTGTTGCCACTGGTGCAATCCAATCCAATGTCCCTGAGGCAATACGCAACTGCTTTGCAGTCTTTCGTACTTTTGCTTGGAACGACAGGATGCCCACGGGAACTTTTCTTGGATCTATATCGCTTCATCCCAACATTAACCCGTACACTGCTCACCTCTCTGGGATGTGGGCCGGGTGGGGCGGTAGTTTTGAGGTCC |
| Astrovirus ^15^ | Forward primer 1 | GGCATGCCTGTTTGACAC |
|  | Forward primer 2 | GGCATGCCTGCTTGACAC |
|  | Reverse primer 1 | ACCTACAGGTTAGTATGACAAC |
|  | Reverse primer 2 | ACCTACAAGTTAGTATGACAAC |
|  | Probe | SUN/ACAGGCACR/ZEN/ACCACGTCATTRTT/3IABkFQ |
|  | Amplicon length | 66 |
|  | Gblock sequence | TCTTGGCATTATCTTCTTGTGCTTCATGGAAGACTCCAACTATGTGAGCCAGATACGTGGCCTTATTGCTACAGCAGTATTAATTGCTGGTGGGCATGCCTGTTTGACACTCACAGGCACGACCACGTCATTGTTTGTTGTCATACTAACCTGTAGGTTCATACGTATGGCAACTGTTTTCATTGGCACCAGGTTCGAGATCCGTGACGCTAATGGAAAGGTTGTGGCCACTGTACCAACTAGGATTAAAAATGTTGCATTTGACTTTTTTCAGAAGCTGAAGCAGTCAGGGGTGCGAGTT |
| Rotavirus A ^15^ | Forward primer | GGACCATCTGATTCTGCTTC |
|  | Reverse primer | GCATTTGTCTTAACTGCATTCG |
|  | Probe | Cy5/CGATCCACT/TAO/CACCAGCTTTTCGATTAG/3IAbRQSp |
|  | Amplicon length | 74 |
|  | Gblock sequence | ATGAATCGTCTTCAACAACGTCAACTCTTTCTGGAAAATCTATTGGTAGGAGTGAACAGTACATTTCACCAGATGCAGAAGCATTCAATAAATACATGCTGTCGAAGTCTCCAGAGGATATTGGACCATCTGATTCTGCTTCAAACGATCCACTCACCAGTTTTTCGATTAGATCGAATGCAGTTAAGACAAATGCAGACGCTGGCGTGTCTATGGATTCATCAGCACAATCACGACCTTCAAGTAATGTCGGATGCGATCAAGTGGATTTCTCCTTAAATAAAGGCTTAAAAGTAAAAGC |
| Adenovirus 41 | Forward primer | AGGCTTCAATAACACAGGAG |
|  | Reverse primer | GAAGAATTAAGTTGGCACCG |
|  | Probe | Cy55/TAAATGCTGCAGGAGGAATGAGAGTG/3IAbRQSp |
|  | Amplicon length | 77 |
|  | Gblock sequence | GGCCACCGCCGCACCACTGACAGTAAGCAACAACCAGCTTAGTATTAACACTGGCAGAGGCTTAGTTATAACTAACAATGCCGTAGCAGTTAATCCTACCGGAGCGTTAGGCTTTAACAACACAGGAGCTTTACAATTAAACGCTGCGGGAGGAATGAGAGTGGACGGCGCCAACTTAATTCTTCATGTAGCATACCCCTTTGAAGCAATCAACCAACTAACACTGCGATTAGAAAACGGGTTAGAAGTAACCAACGGAGGAAAACTCAACGTTAAGTTGGGATCAGGCCTCCAATTTGAC |
| Norovirus GI ^16^ | Forward primer | CGYTGGATGCGNTTYCATGA |
|  | Reverse primer | CTTAGACGCCATCATCATTYAC |
|  | Probe | FAM/TGGACAGGR/ZEN/GAYCGC/3IABkFQ |
|  | Amplicon length | 85 |
| Norovirus GII ^16^ | Forward primer | CARGARBCNATGTTYAGRTGGATGAG |
|  | Reverse primer | TCGACGCCATCTTCATTCACA |
|  | Probe | SUN/TGGGAGGGC/ZEN/GATCGCAATCT/3IABkFQ |
|  | Amplicon length | 98 |
| Norovirus GI + GII | Gblock sequence | GAAATCTACATTCCTGGTTGGCAGGCCATGTTCCGCTGGATGCGATTCCATGATCTAAGTCTGTGGACAGGAGACCGCGATCTCCTGCCCGATTATGTAAATGATGATGGCGTCTAAGGACGCCCCAACAAACATGGATGGCACCAGTGGTGGGGATGGACTTTTACGTGCCAAGGCAGGAACCCATGTTCAGGTGGATGAGGTTTTCTGACTTGAGCACGTGGGAGGGCGATCGCAATCTGGCTCCCAATTTTGTGAATGAAGATGGCGTCGAATGACGCCGCTCCATCTACTGATGGTGCAG |

^*^The source of each ddPCR assays is cited. The ddPCR assays for *Cryptosporidium* and Adenovirus 41 were designed in this study.

**Table SI.7:** Final concentrations of the primer-probe mix for all targets excluding sapovirus and astrovirus

| **Assay component** | **Final concentration (µM)** | **40x concentration (µM)** |
| --- | --- | --- |
| Forward primer | 0.9 | 36 |
| Reverse primer | 0.9 | 36 |
| Probe | 0.25 | 10 |

**Table SI.8:** Final concentrations of the primer-probe mix for sapovirus and astrovirus

| **Assay component** | **Final concentration (µM)** | **40x concentration (µM)** |
| --- | --- | --- |
| Forward primer 1 | 0.45 | 18 |
| Forward primer 2 | 0.45 | 18 |
| Reverse primer 1 | 0.45 | 18 |
| Reverse primer 2 | 0.45 | 18 |
| Probe | 0.25 | 10 |

**Table SI.9:** Reaction composition for RT-ddPCR assay (respiratory and enteric viruses)

| **Reagent** | **Volume (µL)** |
| --- | --- |
| One-step RT-ddPCR supermix | 5.5 |
| Reverse transcript (10x) | 2.2 |
| 300 mM DTT | 1.1 |
| Primer-Probe mix of each target (40x) | 0.55^1^ |
| RNase/DNase-free water | Depending on plate^2^ |
| RNA template | 10 |

**^1^**Final concentrations in reaction: 0.25µM (probe) and 0.9µM (forward and reverse primers)

^2^RNase/DNase-free water for respiratory viruses: 0.45 µL; for enteric viruses (norovirus): 2.1 µL; for enteric viruses (besides norovirus): 1 µL

**Table SI.10:** Reaction composition for ddPCR assay

| **Reagent** | **Volume (µL)** |
| --- | --- |
| ddPCR Multiplex Supermix | 5.5 |
| Primer-Probe mix of each target (40x) | 0.55^*^ |
| RNase/DNase-free water | 3.8 |
| RNA template | 10.5 |

**^*^**Final concentrations in reaction: 0.25µM (probe) and 0.9µM (forward and reverse primers)

**Table SI.11:** Thermal cycling conditions for the respiratory (SARS-CoV-2, influenza A, influenza B, and RSV) and enteric virus (norovirus GI, norovirus GII, sapovirus, astrovirus, rotavirus A, and adenovirus 41) RT-ddPCR assays

| **Cycling step** | **Temperature** °C | **Time** | **Number of cycles** |
| --- | --- | --- | --- |
| Reverse transcription | 50 | 60 min | 1 |
| Enzyme activation | 95 | 10 min | 1 |
| Denaturation | 94 | 30 sec | 40 |
| Annealing/Extension | Depending on plate^*^ | 60 sec |  |
| Enzyme deactivation | 98 | 10 min |  |
| Hold (optional) | 4 | Infinite | 1 |

^*^Annealing temperature for respiratory viruses: 60 °C; for enteric viruses (norovirus): 52.1 °C; for enteric viruses (except norovirus): 59.5 °C

**Table SI.12:** Thermal cycling conditions for pathogenic microorganisms (*Cryptosporidium, Candida auris, Salmonella, and Campylobacter jejuni*) ddPCR assay

| **Cycling step** | **Temperature** °C | **Time** | **Number of cycles** |
| --- | --- | --- | --- |
| Reverse transcription | 50 | 60 min | 1 |
| Enzyme activation | 95 | 10 min | 1 |
| Denaturation | 94 | 30 sec | 40 |
| Annealing/Extension | 60.1 | 60 sec |  |
| Enzyme deactivation | 98 | 10 min |  |
| Hold (optional) | 4 | Infinite | 1 |

#### **SI 1.5 Calculation of recovery rate and inhibition factor**

Recovery rate calculation

**SI. Eq 1:**

$$Recovery rate = \frac{{(C}_{WW-spike}- C_{WW-raw})\times V_{WW}}{C_{std}\times V_{std}}\times100\%$$

Where:

C_ww-spike_: concentration of target DNA or RNA in wastewater spiked with standards using (RT-)ddPCR, copies/L

C_ww-raw_: concentration of target DNA or RNA in raw wastewater using (RT-)ddPCR, copies/L

V_ww_: volume of concentrated wastewater sample, L

C_std_: concentration of standards spiked into wastewater sample, copies/µL

V_std_: volume of standard spiked into each wastewater sample, µL

Inhibition factor calculation

**SI. Eq 2:**

$$Inhibition factor =\frac{C_{10-fold}}{C_{0}}$$

Where:

C_0_: concentration of target DNA or RNA in undiluted extracts, copies/L

C_10-fold_: concentration of target DNA or RNA in10-fold diluted nucleic acid extracts, copies/L

#### **SI 1.6 Quality control measures and the theoretical limit of detection (LOD) calculation**

The quality control measures and the calculation of theoretical LOD are described by Low et al. (2022) ^7^ and Wolken et al. (2023) ^17^ and are summarized briefly as follows. Duplicates of negative control samples were included in the sample treatment, concentration, extraction, and quantification steps to assess potential contamination ^18^. Two aliquots of deionized (DI) water were processed in the same way as the wastewater samples and used as negative controls for concentration. Two bead tubes containing glass beads and lysis buffer were included as extraction negative controls. The negative controls for concentration and extraction were included in all ddPCR quantification plates containing the wastewater samples that were processed together with the controls. In addition, each ddPCR quantification plate included at least two no-template controls (NTCs) with RNAse-free water and two positive controls using gBlock Gene Fragments (IDT, USA; sequence provided in SI 1.3).

An acceptable total droplet count of at least 10,000 was established for all sample wells as recommended by the manufacturer. Each method's theoretical limit of detection (LOD) for ddPCR was determined as three positive droplets per well plus the maximum number of positive droplets among the negative controls. The theoretical LOD was converted to copies per µL of DNA template and copies per liter of wastewater based on the estimated droplet volume (0.86 nL for QX600 and 0.795 nL for QX200), the number of total droplets, the volume fraction of DNA template within a droplet (10/22), and the concentration factor during sample processing. The equations used for the LOD calculation are presented in SI. Eq 3-5.

Theoretical Limit of Detection (LOD) Calculation Equations

**SI. Eq 3:**

$${LOD}_{droplet}=3+maximum number of positive droplets across all process {blanks}^{*}$$

^*^ Process blanks include: two concentration blanks, two extraction blanks, and no less than two no template controls per plate included in ddPCR quantification.

**SI. Eq 4:**

$${LOD}_{\mu L-DNA/RNA template} =\frac{{LOD}_{droplet}}{droplet volume\times n total {droplets in each well}^{*}\times\frac{10}{22}fraction of template within droplet}$$

^*^ The average volume of a droplet is 0.795 nl/droplet for Noroviruses GI and GII (quantified using QX200) and 0.86 nl/droplet for other targets (quantified using QX600)

**SI. Eq 5:**

$${LOD}_{L-wastewater}={LOD}_{\mu L-DNA/RNA template}\times\frac{1}{Concentration factor} \times\frac{1000000 \mu L}{1 L}$$

**Table SI.13:** Summary of concentration factors, equivalent volumes, and theoretical LODs for each concentration method

| **Concentration method** | **Concentration factor** | **Theoretical LOD (copies/L)** | **Equivalent volume** |  |
| --- | --- | --- | --- | --- |
| Direct extraction without bead beating | 6 | 71,586 | 60 | µL |
| Direct extraction with bead beating | 1.8 | 238,622 | 18 | µL |
| HA filtration/solids method | 300 | 1,431 | 3 | mL |
| Nanotrap method without bead beating | 100 | 4,295 | 1 | µL |
| Nanotrap method with bead beating | 30 | 14,317 | 300 | µL |

*The theoretical LOD in copies/L-wastewater was calculated based on a LOD of 0.43 gene copies/µL DNA or RNA template for (RT-)ddPCR. The calculation of LOD used an average droplet count per well of 17,867, which was the lowest among all plates in this study, to provide a conservative estimation.

### **SI 2. Results**

#### **SI 2.1 Examples of *Cryptosporidium* and Adenovirus 41 ddPCR results for samples and controls**

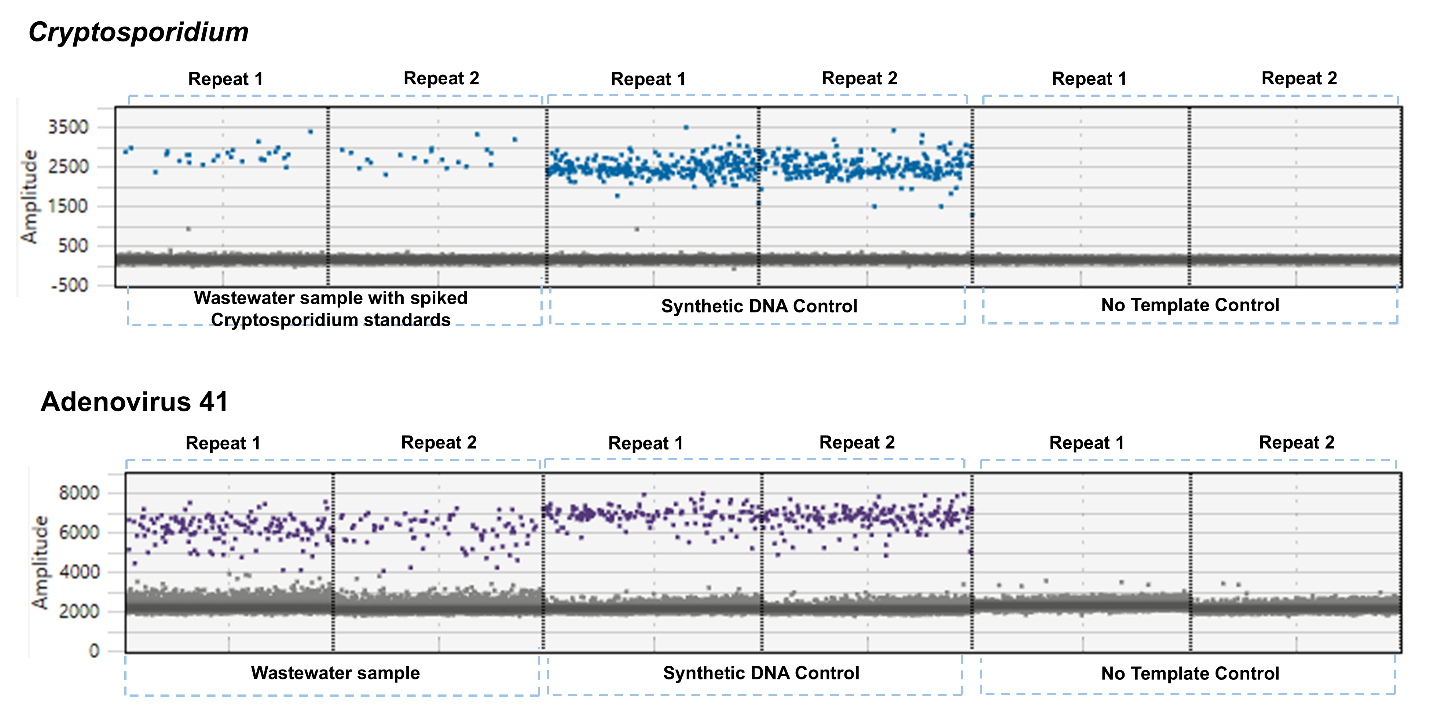

**Figure SI.1** Example plot of samples and control wells for *Cryptosporidium* and Adenovirus 41 ddPCR assays, which were designed in this study. Positive droplets are shown in color based on the target and negative droplets are shown in gray.

#### **SI 2.2 Concentrations of pathogens in spike-in solution differed across quantification methods**

We compared the pathogen concentrations in the spike-in solution determined using three pre-treatment methods (Figure SI.1). For the respiratory viruses, nucleic acid extraction without bead beating yielded the highest concentrations. We observed the highest concentration of *Candida auris* in the spike-in solution when the solution went through bead beating and nucleic acid extraction before being quantified using ddPCR. The highest concentrations for the other target microorganisms were observed using heat-lysis of the solution before quantification.

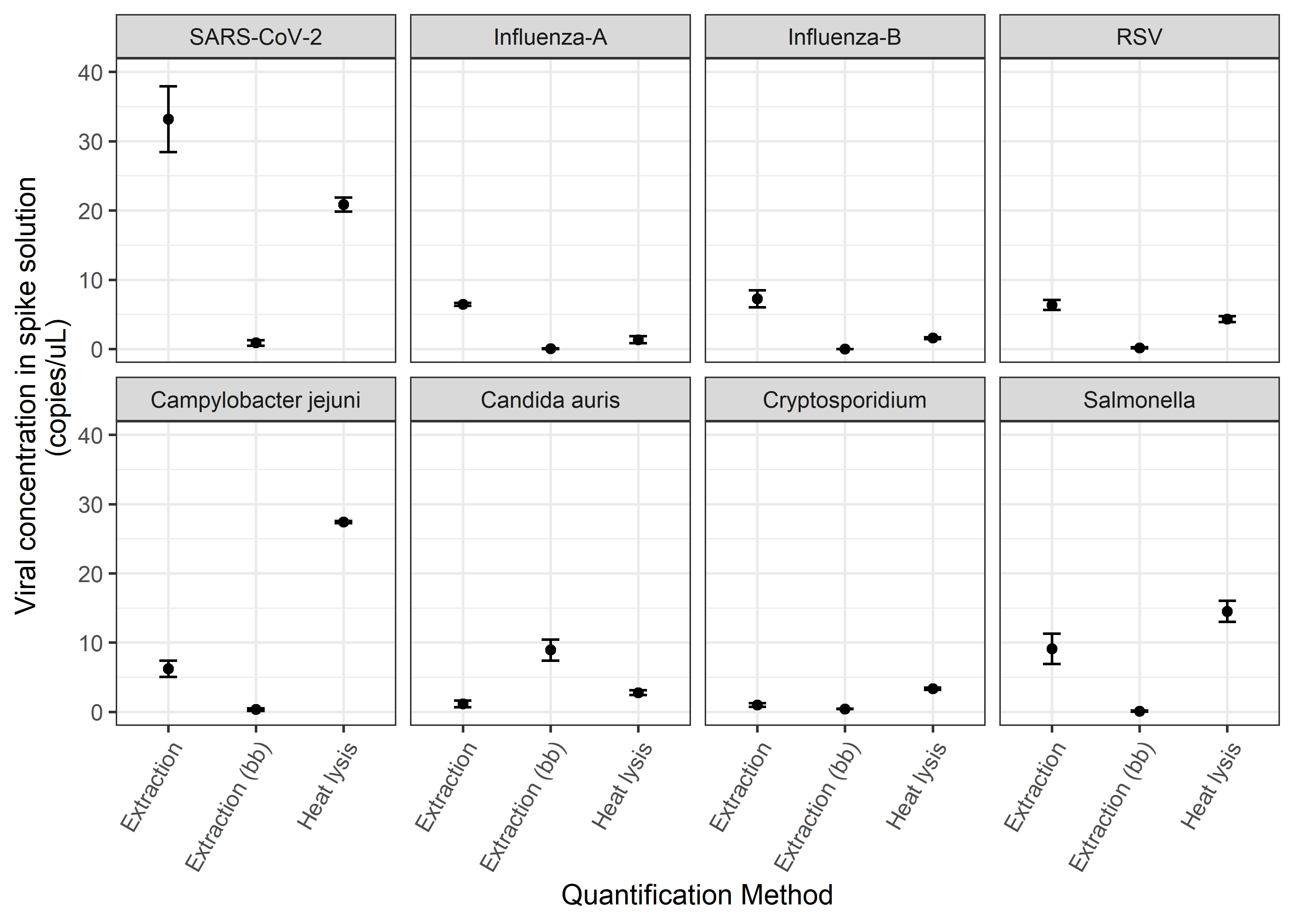
**Figure SI.2.** Impact of quantification method on nucleic acid concentrations measured in the pathogen mixture solution spiked into wastewater samples.

**References:**

(1)	NCBI Virus. *NCBI Virus*. Bethesda (MD): National Library of Medicine (US), National Center for Biotechnology Information. https://www.ncbi.nlm.nih.gov/labs/virus/vssi/#/ (accessed 2025-04-09).

(2)	Untergasser, A.; Cutcutache, I.; Koressaar, T.; Ye, J.; Faircloth, B. C.; Remm, M.; Rozen, S. G. Primer3—New Capabilities and Interfaces. *Nucleic Acids Res.* **2012**, *40* (15), e115. https://doi.org/10.1093/nar/gks596.

(3)	Wang, M. X.; Lou, E. G.; Sapoval, N.; Kim, E.; Kalvapalle, P.; Kille, B.; Elworth, R. A. L.; Liu, Y.; Fu, Y.; Stadler, L. B.; Treangen, T. J. Olivar: Towards Automated Variant Aware Primer Design for Multiplex Tiled Amplicon Sequencing of Pathogens. *Nat. Commun.* **2024**, *15* (1), 6306. https://doi.org/10.1038/s41467-024-49957-9.

(4)	Isaza, J. P.; Galván, A. L.; Polanco, V.; Huang, B.; Matveyev, A. V.; Serrano, M. G.; Manque, P.; Buck, G. A.; Alzate, J. F. Revisiting the Reference Genomes of Human Pathogenic Cryptosporidium Species: Reannotation of C. Parvum Iowa and a New C. Hominis Reference. *Sci. Rep.* **2015**, *5* (1), 16324. https://doi.org/10.1038/srep16324.

(5)	Altschul, S. F.; Gish, W.; Miller, W.; Myers, E. W.; Lipman, D. J. Basic Local Alignment Search Tool. *J. Mol. Biol.* **1990**, *215* (3), 403–410. https://doi.org/10.1016/S0022-2836(05)80360-2.

(6)	Laturner, Z. W.; Zong, D. M.; Kalvapalle, P.; Gamas, K. R.; Terwilliger, A.; Crosby, T.; Ali, P.; Avadhanula, V.; Santos, H. H.; Weesner, K.; Hopkins, L.; Piedra, P. A.; Maresso, A. W.; Stadler, L. B. Evaluating Recovery, Cost, and Throughput of Different Concentration Methods for SARS-CoV-2 Wastewater-Based Epidemiology. *Water Res.* **2021**, *197*, 117043. https://doi.org/10.1016/j.watres.2021.117043.

(7)	Lou, E. G.; Sapoval, N.; McCall, C.; Bauhs, L.; Carlson-Stadler, R.; Kalvapalle, P.; Lai, Y.; Palmer, K.; Penn, R.; Rich, W.; Wolken, M.; Brown, P.; Ensor, K. B.; Hopkins, L.; Treangen, T. J.; Stadler, L. B. Direct Comparison of RT-ddPCR and Targeted Amplicon Sequencing for SARS-CoV-2 Mutation Monitoring in Wastewater. *Sci. Total Environ.* **2022**, *833*, 155059. https://doi.org/10.1016/j.scitotenv.2022.155059.

(8)	CDC. *Labs*. Centers for Disease Control and Prevention. https://www.cdc.gov/coronavirus/2019-ncov/lab/rt-pcr-panel-primer-probes.html (accessed 2022-08-17).

(9)	Whiley, D. M.; Sloots, T. P. A 5′-Nuclease Real-Time Reverse Transcriptase–Polymerase Chain Reaction Assay for the Detection of a Broad Range of Influenza A Subtypes, Including H5N1. *Diagn. Microbiol. Infect. Dis.* **2005**, *53* (4), 335–337. https://doi.org/10.1016/j.diagmicrobio.2005.08.002.

(10)	van Elden, L. J. R.; Nijhuis, M.; Schipper, P.; Schuurman, R.; van Loon, A. M. Simultaneous Detection of Influenza Viruses A and B Using Real-Time Quantitative PCR. *J. Clin. Microbiol.* **2001**, *39* (1), 196–200. https://doi.org/10.1128/JCM.39.1.196-200.2001.

(11)	Hughes, B.; Duong, D.; White, B. J.; Wigginton, K. R.; Chan, E. M. G.; Wolfe, M. K.; Boehm, A. B. Respiratory Syncytial Virus (RSV) RNA in Wastewater Settled Solids Reflects RSV Clinical Positivity Rates. *Environ. Sci. Technol. Lett.* **2022**, *9* (2), 173–178. https://doi.org/10.1021/acs.estlett.1c00963.

(12)	Leach, L.; Zhu, Y.; Chaturvedi, S. Development and Validation of a Real-Time PCR Assay for Rapid Detection of Candida Auris from Surveillance Samples. *J. Clin. Microbiol.* **2018**, *56* (2), 10.1128/jcm.01223-17. https://doi.org/10.1128/jcm.01223-17.

(13)	Kasturi, K. N.; Drgon, T. Real-Time PCR Method for Detection of Salmonella Spp. in Environmental Samples. *Appl. Environ. Microbiol.* **2017**, *83* (14), e00644-17. https://doi.org/10.1128/AEM.00644-17.

(14)	Liu, K. C.; Jinneman, K. C.; Neal-McKinney, J.; Wu, W.-H.; Rice, D. H. Simultaneous Identification of Campylobacter Jejuni, Campylobacter Coli, and Campylobacter Lari with SmartCycler-Based Multiplex Quantitative Polymerase Chain Reaction. *Foodborne Pathog. Dis.* **2017**, *14* (7), 371–378. https://doi.org/10.1089/fpd.2016.2245.

(15)	Wolken, M.; Wang, M.; Schedler, J.; Campos, R. H.; Ensor, K.; Hopkins, L.; Treangen, T.; Stadler, L. B. PreK-12 School and Citywide Wastewater Monitoring of the Enteric Viruses Astrovirus, Rotavirus, and Sapovirus. *Sci. Total Environ.* **2024**, *931*, 172683. https://doi.org/10.1016/j.scitotenv.2024.172683.

(16)	Kageyama, T.; Kojima, S.; Shinohara, M.; Uchida, K.; Fukushi, S.; Hoshino, F. B.; Takeda, N.; Katayama, K. Broadly Reactive and Highly Sensitive Assay for Norwalk-like Viruses Based on Real-Time Quantitative Reverse Transcription-PCR. *J. Clin. Microbiol.* **2003**, *41* (4), 1548–1557. https://doi.org/10.1128/JCM.41.4.1548-1557.2003.

(17)	Wolken, M.; Sun, T.; McCall, C.; Schneider, R.; Caton, K.; Hundley, C.; Hopkins, L.; Ensor, K.; Domakonda, K.; Kalvapalle, P.; Persse, D.; Williams, S.; Stadler, L. B. Wastewater Surveillance of SARS-CoV-2 and Influenza in preK-12 Schools Shows School, Community, and Citywide Infections. *Water Res.* **2023**, *231*, 119648. https://doi.org/10.1016/j.watres.2023.119648.

(18)	Borchardt, M. A.; Boehm, A. B.; Salit, M.; Spencer, S. K.; Wigginton, K. R.; Noble, R. T. The Environmental Microbiology Minimum Information (EMMI) Guidelines: qPCR and dPCR Quality and Reporting for Environmental Microbiology. *Environ. Sci. Technol.* **2021**, *55* (15), 10210–10223. https://doi.org/10.1021/acs.est.1c01767.
